# Genetics of cannabis ever-use and frequency across ancestries implicate novel loci and brain-specific biology

**DOI:** 10.64898/2026.04.25.26351611

**Authors:** Joëlle A. Pasman, Zachary F. Gerring, Jackson Thorp, Abdel Abdellaoui, Anil Ori, Pierre Youssef, Muhannad Smadi, Anaïs B. Thijssen, Damian Woodward, Briar Wormington, Daniel E. Adkins, Fazil Aliev, Chris Chatzinakos, Sarah L. Elson, Pierre Fontanillas, Ian R. Gizer, Haixia Gu, Lindsey A. Hines, Emma C. Johnson, Kadri Kõiv, Penelope A. Lind, Miriam A Mosing, Ilja M. Nolte, Jue-Sheng Ong, Jacqueline M. Otto, Teemu Palviainen, Roseann E. Peterson, Hannah M. Sallis, Andrey A. Shabalin, Jean Shin, Nathaniel S. Thomas, Camiel M. van der Laan, Peter J. van der Most, Saskia van Dorsselaer, Kristel R. van Eijk, Robyn E. Wootton, Stephanie Zellers, Catharina A. Hartman, Georgi Hudjashov, Marco P. Boks, Dorret I. Boomsma, Enda M. Byrne, William E. Copeland, Danielle M. Dick, Bart Ferweda, Andrew C. Heath, Ian B. Hickie, William G. Iacono, Martin A. Kennedy, Kelli Lehto, Anja Lok, Stuart MacGregor, Pamela A.F. Madden, Hermine H.M. Maes, Nick G. Martin, Matt McGue, Sarah E. Medland, Marcus R. Munafò, Max Nieuwdorp, Albertine J. Oldehinkel, Miina Ollikainen, Abraham A. Palmer, Tomas Paus, Zdenka Pausova, John F. Pearson, Sandra Sanchez-Roige, Harold Snieder, Margreet ten Have, Jorien L. Treur, Scott Vrieze, Kirk C. Wilhelmsen, Aeilko H. Zwinderman, Estonian Biobank Research Team, Lifelines Cohort Study, International Cannabis Consortium, Jacqueline M. Vink, Nathan Gillespie, Eske M. Derks, Karin J.H. Verweij

## Abstract

Cannabis use is widespread, with genetic differences partly explaining variation in individual patterns of use. We performed the largest-to-date genome-wide association study (GWAS) meta-analysis of cannabis ever-use (N=736,322, 76% European ancestry) and various measures of frequency of use (N=269,160 cannabis users, 84% European ancestry). We identified 54 independent genome-wide significant loci for ever-use and 6 for frequency and show that the genetic architecture of ever-use, frequency, and cannabis use disorder (CUD) are overlapping but distinguishable. We identified 63 loci that were associated with common liability (‘*All-cannabis*’) to different cannabis use traits in European-ancestry individuals. Across analyses, we identified 75 unique loci that had not previously been implicated in cannabis use. Gene prioritization analyses identified 349 genes for ever-use, 5 genes for frequency of use, and 429 for *All-cannabis*, including previously identified and novel genes. We found enrichment of genetic signals for cannabis use in biologically meaningful categories and relevant human brain cell types, including excitatory neuronal populations. There were substantial genetic correlations between cannabis use and a range of psychiatric disorders and substance use traits, while cannabis polygenic scores were associated with increased risk of psychiatric disorders. Mendelian Randomization showed evidence for (bidirectional) causal associations between cannabis use and ADHD, bipolar disorder, schizophrenia and PTSD.

## Introduction

Cannabis is one of the most commonly used psychoactive substances^1^. Twin studies have shown moderate heritability estimates for ever-use (i.e., using at least once during lifetime) and frequency of cannabis use (40-74%) and somewhat higher heritability estimates for measures of Cannabis Use Disorder (CUD; 51-78%)^2,3^. Several GWASs have been conducted to map the genetic architecture underlying cannabis use traits (e.g. ^4-8^). The largest GWAS of cannabis ever-use^9^ (N=271,134) identified 11 loci, with top variants residing in the *CADM2* cell adhesion molecule gene, which has been widely implicated in mental health and risk behaviors^10-13^. For CUD, a GWAS has identified 22 independent loci in individuals of European ancestry, 2 loci each in African and East Asian ancestry, and 1 in admixed American ancestry (total N=1,054,365; 64,314 cases), with a key association with variants in the *CHRNA2* nicotinic acetylcholine receptor gene^6^. These findings represented a significant step forward in terms of identifying genetic variants associated with cannabis use traits across broad ancestral populations.

Ever-use of cannabis and CUD are binary measures, with ever use capturing anything from single-time use to dependence, whereas CUD captures only severe forms of use. Few well-powered GWASs have examined continuous, more granular measures of cannabis use, such as frequency of use, and none have done so across diverse ancestral populations to enhance generalizability. In this study, we present a GWAS of cannabis use frequency, using a meta-analytic sample of 269,160 cannabis users, including 42,163 individuals of American (Am), South Asian (S), East Asian (E), African (Af), and other (O) ancestries (abbreviated throughout as *AmSEAfO*). We also update the GWAS meta-analysis of ever-use, almost tripling the sample size of the previous largest GWAS^9^ to N=736,322, including N=179,862 of *AmSEAfO* ancestry. We contrast and combine measures of different cannabis use traits, including the latest GWAS of CUD^6^, and compare associations with other traits. Specifically, we leverage the GWAS meta-analysis summary statistics to perform secondary analyses, including positional and functional fine-mapping of SNPs and genes, protein and transcriptome-wide association analyses, single-cell enrichment, genetic correlations with a range of other traits, polygenic score analyses, and use Mendelian Randomization (MR) analyses to explore causal relations. This work provides the largest genetic characterization of cannabis use to date and shows that ever-use, frequency, and CUD reflect overlapping but partly distinct genetic architectures, refining how liability across the cannabis use spectrum is conceptualized and informing biological mechanisms and links to psychiatric disorders.

## Results

### GWAS meta-analyses identify 54 and 6 significant loci for ever-use and frequency of use

An overview of the analyses is given in <u>Figure 0</u>, with results summarized in <u>Table S2</u>. GWAS meta-analyses were performed on ever-use of cannabis (comparing those who had used cannabis at least once during their lifetime versus those who never-used cannabis) as well as three measures of frequency of use: lifetime frequency (number of times used during lifetime), maximum frequency (frequency of use during period that the individual used most often), and recent frequency (frequency of use in the past period (e.g., months/year)).

The trans-ancestry meta-analysis (European plus *AmSEAfO* ancestries) of cannabis ever-use identified 55 independent genome-wide significant loci, with the strongest locus on chromosome 3 in *CADM2* (Figure 1A, <u>Table S3</u>). All SNPs combined accounted for 5.4% of the variance in ever-use (SNP-based heritability h^2^_SNP_ =5.4%, standard error SE=0.3%). There were 47 loci novel loci compared to a previous cannabis ever-use GWAS and 47 compared to CUD (<u>Table S4-S5</u>). The European-ancestry-only GWAS identified 45 significant loci (h^2^_SNP_ =6.5%, SE=0.3%), including the same lead locus in *CADM2*. Nine European loci did not reach genome-wide significance in the trans-ancestry GWAS, including two intergenic loci on chromosome X (Figure 1A, <u>Table S3</u>). On the other hand, 15 loci from the trans-ancestry GWAS were not significant in the European-only GWAS. GWAS results excluding European individuals yielded 11 significant loci and are summarized in <u>Figure S1</u> and <u>Table S3</u>. The genetic correlation between the European and *AmSEAfO* results was significantly different from 1, r_g=_0.62 (SE=0.06, p=5.04e-12). None of the 11 loci significant in the *AmSEAfO* GWAS were among the trans-ancestry or European hits. We queried the lead SNP in each locus that was uniquely significant in the trans-ancestry, European, and/or *AmSEAfO* analysis in GWASAtlas (66 SNPs), revealing associations with alcohol use and tobacco smoking behaviors, schizophrenia, and educational attainment, while 18 SNPs had no previous associations (<u>Table S6</u>).

**Figure 1.**
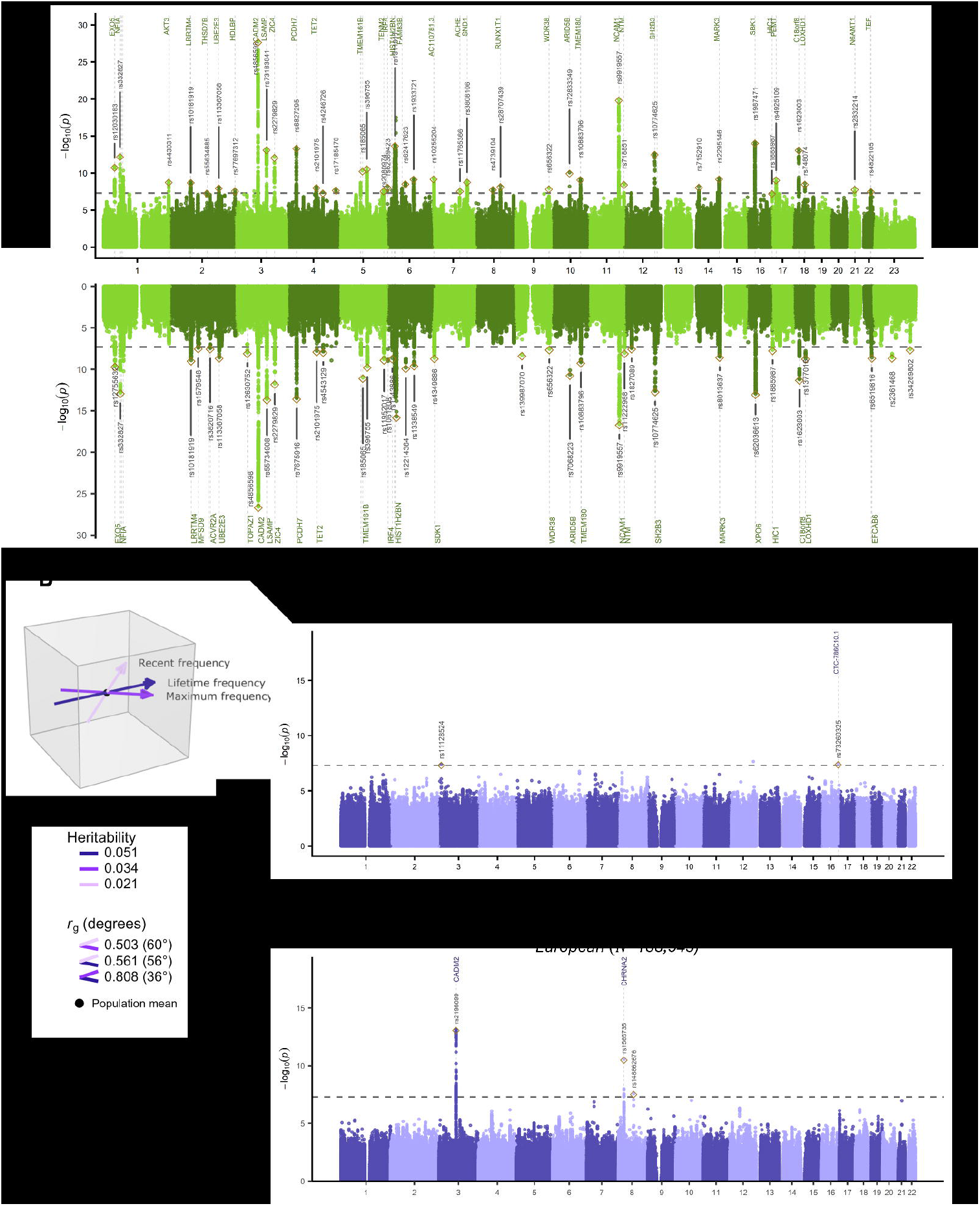
GWAS results. **A)** Manhattan plots of the p-values of each SNP association with ever-use across different ancestries (top) and in European-ancestry only (bottom). **B)** Visualization of the genetic relationship between the 3 frequency traits, with smaller angles representing higher genetic correlations and the length of the arrows the heritabilities (interactive 3D version in <u>Figure S4</u>). **C)** Manhattan plot of the recent frequency variable across different ancestries. **D)** Manhattan plot of the latent *All-frequency* variable in European-ancestry individuals (combining lifetime, maximum, and recent frequency of use). **A/C**: If SNPs could be positionally mapped to their nearest genes, gene names are displayed. Top SNPs in each locus are annotated (pruning some overlapping labels); the full list of top SNPs can be found in <u>Table S3</u> (ever-use), <u>S9</u> (recent frequency), and <u>S11</u> (*All-frequency*). The horizontal dashed lines indicate the genome-wide significance threshold of *p*<5e-8.

European-ancestry GWASs for the three measures of cannabis use frequency identified i) 2 genome-wide significant loci for lifetime frequency (h^2^_SNP_ =5.1%, SE=0.9%), ii) 2 loci in the same regions for maximum frequency (h^2^_SNP_ =3.4%, SE=0.5%), and iii) none for recent frequency (h^2^_SNP_ =2.1%, SE=0.6%; <u>Figure S2</u>; <u>Table S7-S8</u>). Of the 3 unique loci identified for cannabis use frequency, 1 was unique compared to the ever-use GWAS, namely a locus in the GULOP gene. The trans-ancestry meta-analysis of recent frequency (which was the only available frequency phenotype for *AmSEAfO* ancestries) identified 2 other genome-wide significant loci (h^2^_SNP_ =1.9%, SE=0.3%; N=128,116, 32.9% *AmSEAfO*; Figure 1C, <u>Table S9</u>, with the results excluding European-ancestry individuals separately in <u>Figure S3</u> and <u>Table S10</u>). The correlation between the European and AmSEAfO recent frequency GWAS was not significantly different from 1, r_g=_0.96 (SE=0.32, p=0.897).

### Combining frequency traits identifies an additional risk locus

The three European-ancestry frequency GWASs exhibited overlapping genetic architectures, as shown by the genetic correlations (ranging between r_g_=0.50-0.81; only estimated in the European sample due to lack of reference data with the same ancestry composition as our GWAS) as well as a visualization of their relation in GDIS (Figure 1B). The small angles between the traits indicate strong genetic correlations and the similar lengths of their vectors reflect comparable SNP-heritability estimates. We therefore applied Genomic SEM to model a common latent *All-frequency* factor that integrates these three European-only GWASs (analytical sample size 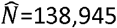); this factor identified 3 independent genome-wide significant loci (1 extra with respect to the separate frequency GWASs: counting the loci from the trans-ancestry GWAS the number of loci for frequency now totals 6; h^2^_SNP=_4.1%, SE=0.5%; Figure 1D, <u>Table S11</u>). Two loci were novel compared to the previously published ever-use GWAS (<u>Table S12</u>), and 1 was also novel compared to the previous CUD GWAS (<u>Table S13</u>) and to GWASs of other traits in GWASAtlas (<u>Table S14</u>). Across all GWASs (trans-ancestry ever-use and recent frequency, European-only *All-frequency* and *All-cannabis*) 75 independent loci were novel compared to the previous ever-use and CUD GWASs.

### Shared and distinct genetic liability to different stages of cannabis use in European ancestry

Continuing with the European-ancestry GWAS results, we mapped the shared and distinct genetic liability to different cannabis use traits. First, we estimated genetic correlations between ever-use, *All-frequency*, and CUD, finding moderate to high overlap across traits (*r*_*g*_=0.60-0.76). We then used Genomic Network Analysis (GNA) to estimate partial genetic correlations between the three traits (that is, the genetic correlation between traits when partialling out the effects of the third trait). We found that the genetic correlation between CUD and ever-use (r_g_=0.60, p=6.94e-63) was not significant after accounting for *All-frequency* (partial *r*_*g*_=0.08, p=0.547; Figure 2A, <u>Table S15</u>). Thus, the overlap between ever-use and CUD was entirely explained by *All-frequency*. To validate this, we fitted a sparse network model in GNA in which the edge between CUD and ever-use was fixed to 0 and estimated the remaining two edges. The model fit was excellent (Standardized Root Mean Square Residual [SRMR]=0.013, Comparative Fit Index [CFI]=0.999), thereby supporting *All-frequency* mediating the genetic correlation between CUD and ever-use (<u>Table S16</u>).

**Figure 2.**
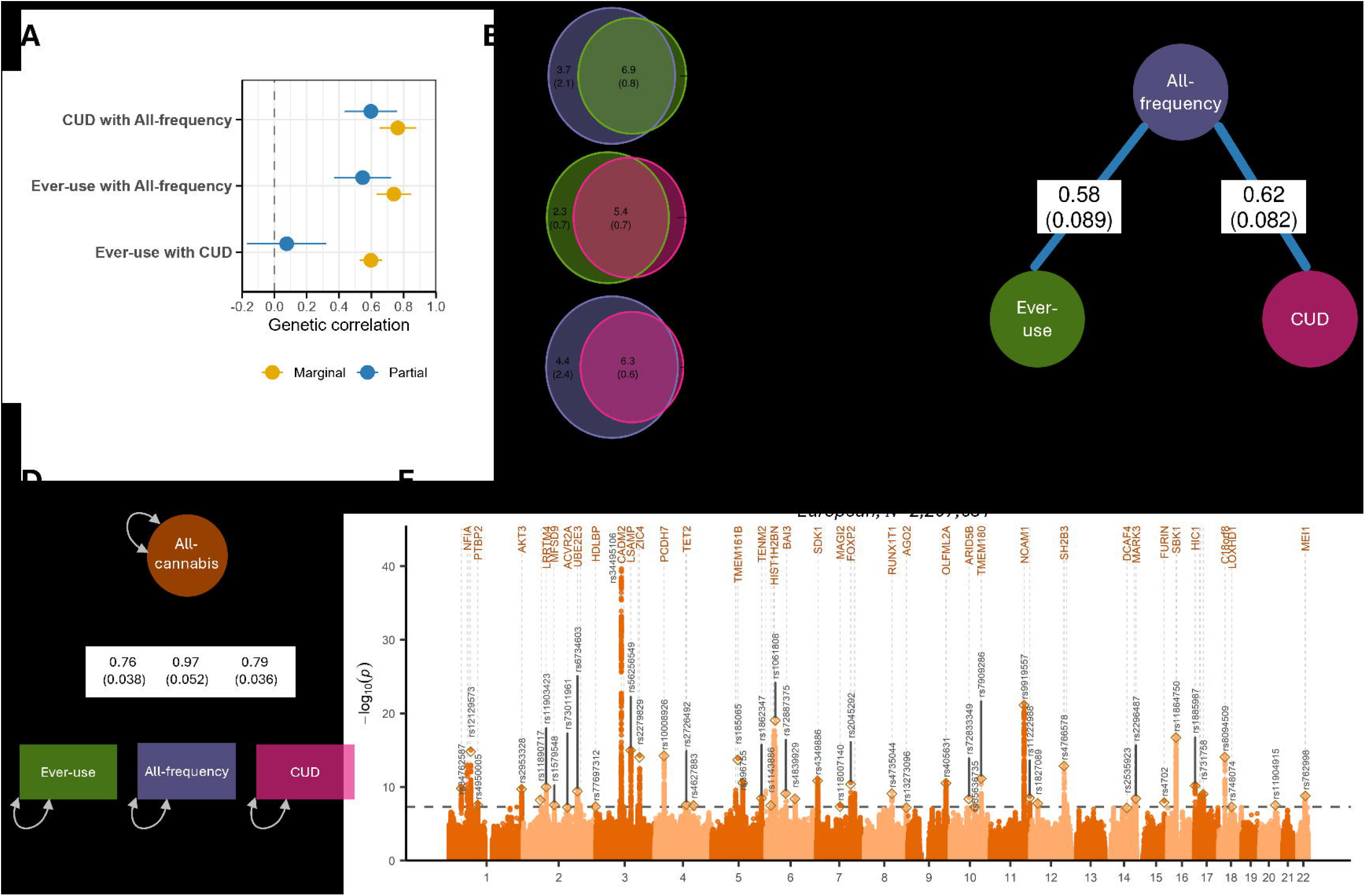
Common genetic liability to cannabis use behavior. **A)** Bivariate genetic correlations (yellow) as estimated with LD Score Regression, and partial genetic correlations (blue) estimated with Genomic Network Analysis. Error bars are 95% confidence intervals. Full results are in <u>Table S14</u>. **B)** Polygenic overlap as estimated with MiXeR; the number of causal variants that are unique or shared between traits. Numbers represent thousands of variants (SE). **C)** Results from a sparse network model estimated with Genomic Network Analysis, with edges representing the partial genetic correlations between traits (with the non-significant edge between ever-use and CUD pruned from the network). **D)** Path diagram of common factor model estimated with Genomic structural equation modelling. **E)** Manhattan plot of SNP effects from the multivariate GWAS of the *All-cannabis* latent factor.

We next applied MiXeR to characterize the polygenicity of the cannabis use traits in European-ancestry individuals by estimating the number of causal variants influencing each trait and their polygenic overlap. *All-frequency* was the most polygenic (10.6k causal variants, SE⍰=⍰2.5k), followed by ever-use (7.7k causal variants, SE⍰= ⍰0.66k), whereas CUD was the least polygenic (6.5k causal variants, SE⍰= ⍰0.67k). We then estimated the number of shared and unique causal variants for each trait. The vast majority were shared (Figure 2B; <u>Table S17</u>). Furthermore, for all three trait pairs, polygenic overlap was best explained by a model in which the causal variants of the least polygenic trait formed a subset of the causal variants in the most polygenic trait: all variants that influence CUD also influence ever-use, and all variants that influence ever-use also influence *All-frequency*.

Given the considerable polygenic overlap between *All-frequency*, ever-use and CUD, we fitted a common factor model in order to capture genetic effects that are shared between the three cannabis traits (Figure 2D, <u>Table S18</u>). Multivariate GWAS of this latent factor (*All-cannabis*) identified 63 independent genome-wide significant loci (57 novel loci) but resulted in a lower SNP-based heritability (estimated 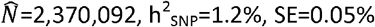, <u>Table S19-S22</u>).

### Novel genes identified for cannabis use

Gene-based association analyses were performed on the summary statistics from three GWASs: (1) the latent *All-frequency* factor; (2) ever-use; and (3) the *All-cannabis* factor. Significant genes (Bonferroni-corrected p<.05) were classified into three evidence tiers: Tier 1 (high-confidence genes) required converging support from proximity-based tests (MAGMA or mBAT) and at least one expression-based test of association (TWAS with COLOC validation or SMR with HEIDI validation); Tier 2 genes either showed significant proximity-based tests alone or multiple independent expression-based signals without full multimodal support; and Tier 3 genes comprised genes significant in only a single test type.

Across all three analyses, we identified 446 significant unique genes in at least one gene-based test (summarized in <u>Table S23-S25</u>; full results <u>Table S26-S43</u>). The *All-cannabis* factor yielded the most associations (429 genes: 33 Tier 1; 62 Tier 2; 334 Tier 3 genes), followed by ever-use (349 genes: 22 Tier 1; 52 Tier 2; 275 Tier 3) and the *All-frequency* factor (5 genes: 1 Tier 1; 1 Tier 2; 3 Tier 3). Focusing on the high-confidence Tier 1 genes, the *All-cannabis* factor and ever-use had substantial overlap: of the 22 Tier 1 genes for ever-use and 33 for *All-cannabis*, 19 genes were prioritized in both (Figure 3A). When expanding to all three traits, we identified 37 unique Tier 1 genes in total. No single gene met Tier 1 criteria across all three traits (Figure 3B).

**Figure 3.**
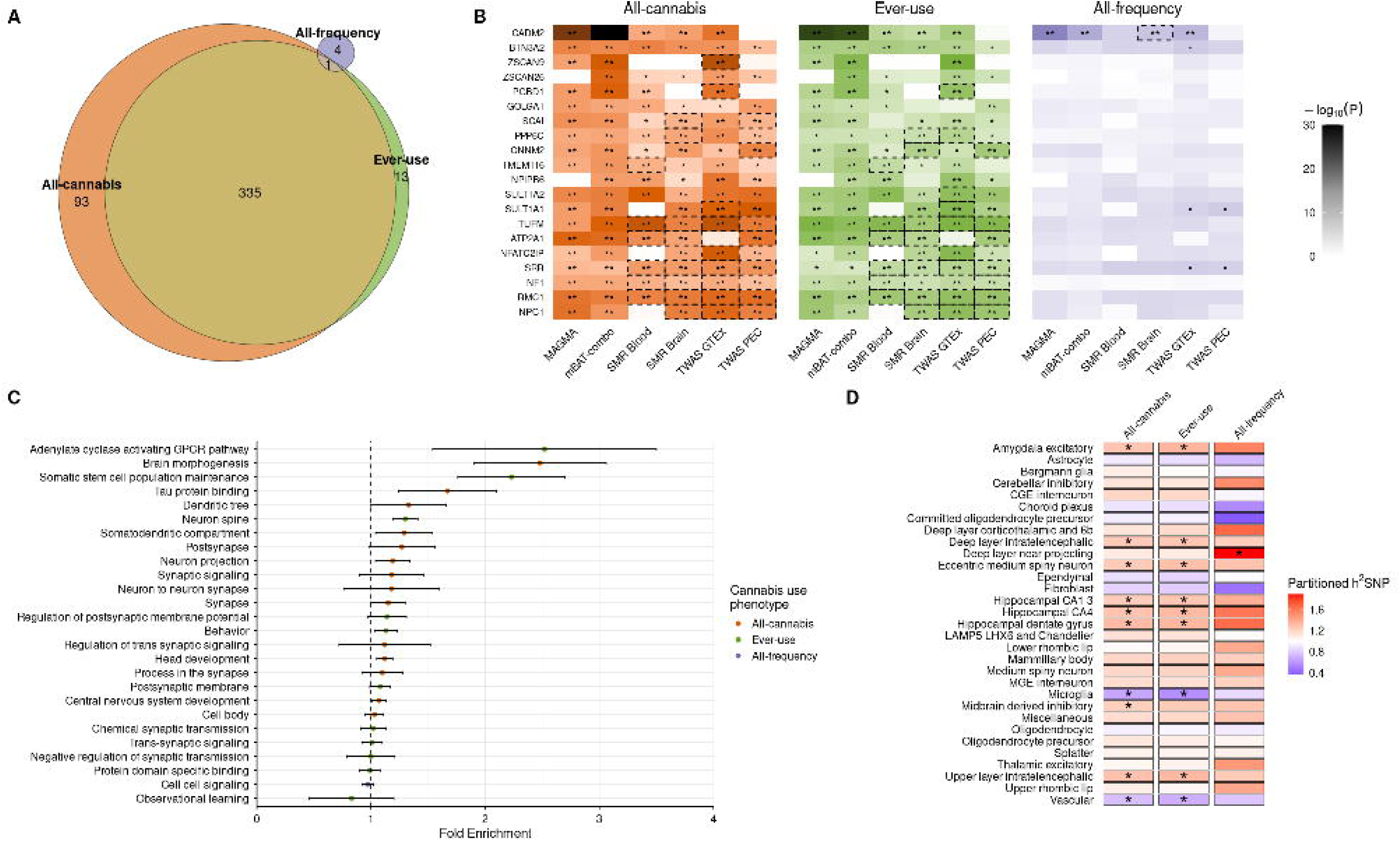
High confidence genes and functional annotation. **A)** Venn diagram of the number of unique significant (Bonferroni-corrected p-value *p*_*Bonferroni*_<0.05) genes (i.e., Tier 3 genes) from six gene-based tests for each cannabis use trait. **B)** Top 20 Tier 1 and Tier 2 genes (based on mean -log10(*p*-value) across each phenotype) ordered by chromosomal location. **C)** Gene sets prioritized by MAGMA (*p*_*Bonferroni*_<0.05), ranked by GSA-MiXeR fold enrichment. GSA MiXeR fold enrichment estimates with standard errors; the vertical dashed line denotes enrichment threshold (fold enrichment=1). **D)** SNP-heritability enrichment in 31 human brain cell type superclusters for 3 cannabis traits. *False discovery rate-corrected *p*_*FDR*_<0.05; \*\**p*_*Bonferroni*_<0.05; dashed boxes highlight gene-based associations with colocalisation posterior probability >0.8 (TWAS) or HEIDI *p*>0.0.5 (SMR).

A look-up in GWAS catalog showed that all of the Top 20 Tier 1 and 2 genes have previously been identified in relation to at least 4 traits, with CADM2 previously associated with the greatest number of traits (284; <u>Table S44</u>). Thirteen of these 20 genes have been previously identified in relation to any substance use trait, three of which (*CADM2, PPP6C, ATP2A1*) have been previously linked to a cannabis use trait. The three genes in the 6p22.1 locus have not been previously associated with substance use traits but have been identified in relation to other psychiatric traits such as major depressive disorder, autism spectrum disorder, or schizophrenia. Seven genes have also been previously identified in relation to risk-taking/externalizing behavior, which is known to correlate with cannabis use phenotypically and genetically. Other traits commonly associated with these 20 genes appear to be related to body mass index, body measurements (e.g. height, waist, and hip circumference), or blood serum measurements (e.g., cholesterol and platelet count) among many others.

### Cannabis use enriched for neuronal signaling pathways

We next applied GSA-MiXeR to detect polygenic enrichment in curated gene sets for each of the three GWASs (ever-use, *All-frequency*, and *All-cannabis*) separately (Figure 3C, <u>Table S45</u>). Significant (Bonferroni-corrected) enrichments related to “adenylate cyclase activating G protein coupled receptor signaling” pathways, “neuron spine,” and “behavior” were observed in the ever-use phenotype, while gene sets related to brain morphogenesis and tau protein binding and neurons were enriched in genetic signal for the *All-cannabis* factor.

Partitioned heritability analyses showed broadly consistent cell-type-specific patterns for ever-use and *All-cannabis* (Figure 3D; <u>Table S46</u>). For example, both traits had significant enrichment in excitatory neuronal populations—most notably amygdala excitatory neurons, hippocampal neurons, and intratelencephalic neurons. We also identified significant depletion of genetic signal for ever-use and *All-frequency* in vascular cells and microglia, indicating a predominantly neuronal rather than glial or vascular contribution to genetic risk. In contrast, *All-frequency* displayed a distinct profile, with its strongest and most specific enrichment observed in deep-layer near-projecting neurons, a signal not detected for ever-use or *All-cannabis*. These results suggest partially shared but also phenotype-specific cellular architectures underlying the different cannabis use-related traits.

### Genetic liability to stages of cannabis use shows partially distinct associations with external traits

Genetic correlations with other complex traits (Figure 4, <u>Table S47</u>) were estimated for ever-use, *All-frequency*, and CUD^6^, to examine potential differences across the cannabis use spectrum. Interestingly, ever-use showed modest positive correlations with (favorable) cognitive and socioeconomic outcomes (e.g., educational attainment, income, and IQ), whereas CUD revealed substantial negative correlations. *All-frequency* had near-zero or very weak correlations with these traits. All cannabis measures showed moderate-to-strong positive genetic correlations with the Townsend deprivation index and weak positive correlations with working memory. They were genetically associated with most mental health and substance use traits in the risk-increasing direction, with associations generally strongest for CUD, followed by frequency of use, and weakest for ever-use. Genetic correlations of all cannabis traits with personality traits showed negative associations with conscientiousness and positive associations with openness to experience as well as extraversion, whereas ever-use and CUD revealed negative associations with neuroticism.

**Figure 4.**
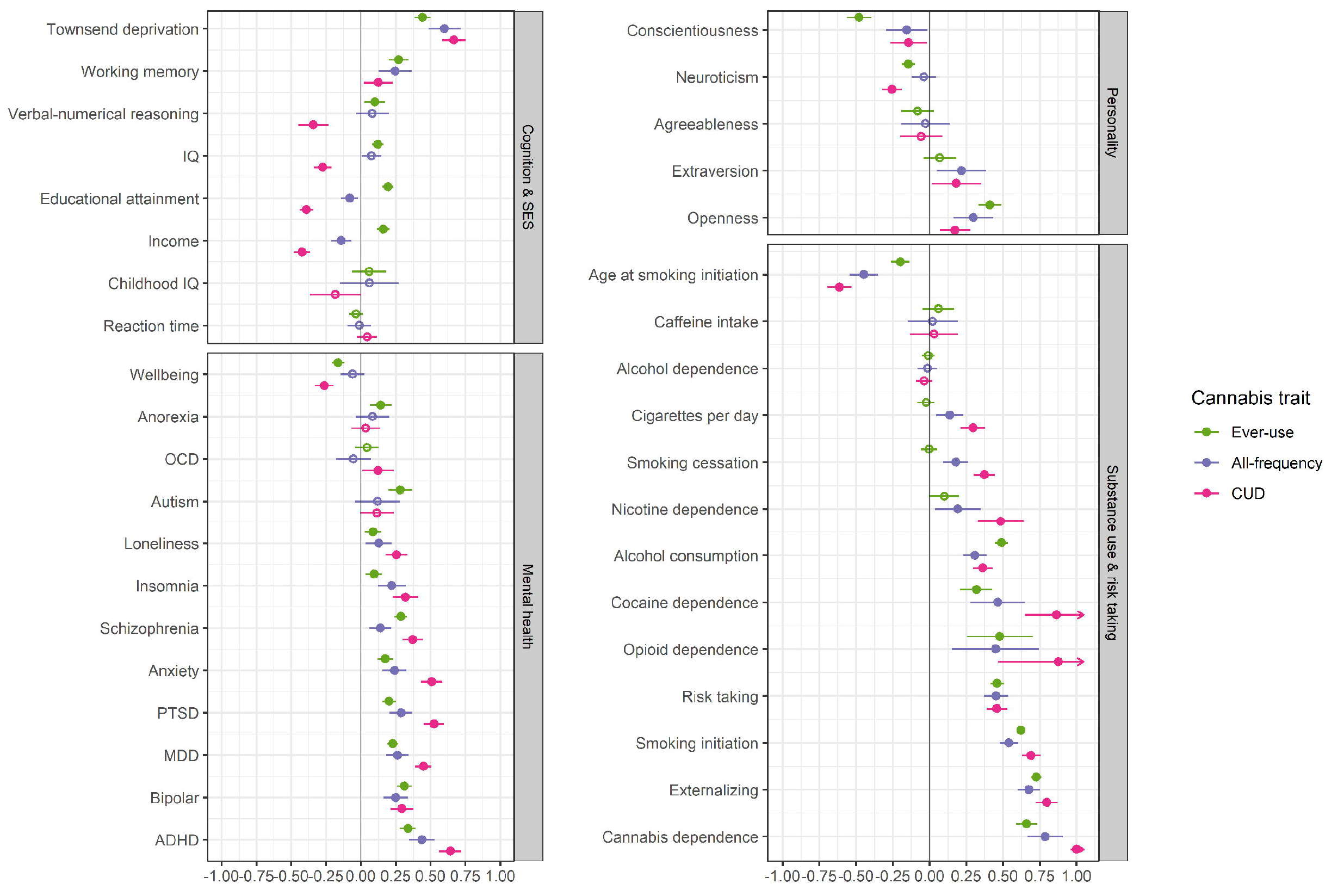
Genetic correlations between cannabis use and external traits with 95% CI. Filled circles are significant at *p*_FDR_<0.05. Trait sources are described in <u>Table S48</u>. *MDD = major depressive disorder; OCD = obsessive-compulsive disorder; PTSD = posttraumatic stress disorder; IQ = intelligence quotient; ADHD = attention deficit/hyperactivity disorder*

**Figure 5.**
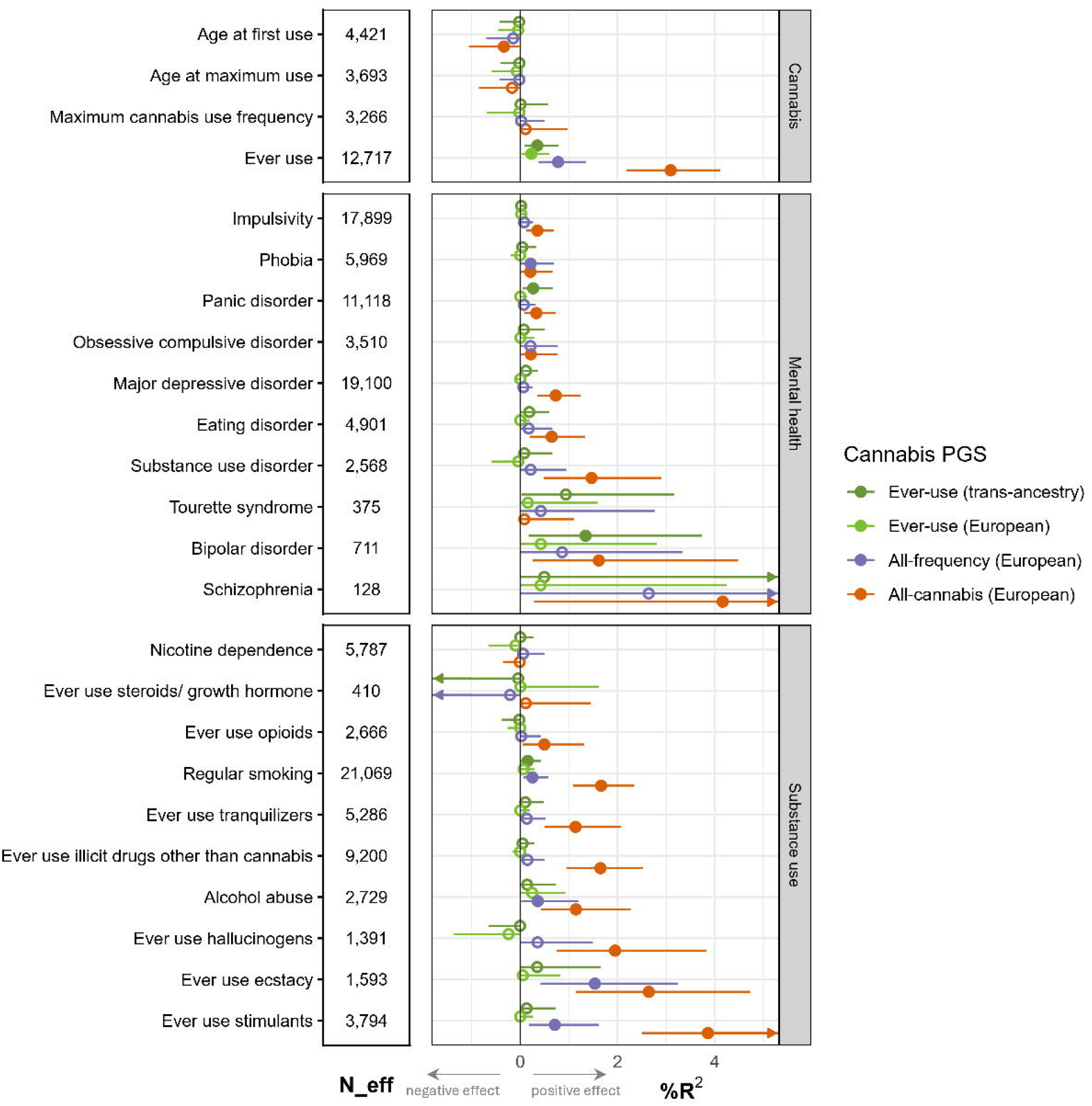
Results from the polygenic score analysis in the STAGE sample, with corresponding effective sample size (N_eff). Outcome measures are described in <u>Table S50</u>. The circles denote the percentage explained variance (%R^2^ ) by the PGS in each trait, with the asymmetrical CI acquired with bootstrapping. %R^2^ and CI were converted to a negative estimate when 11<0 for visualization purposes. For binary traits, Nagelkerke’s R^2^ was used. Filled circles signify significant R^2^ (i.e., model with PGS explained significantly more variance than model without PGS) after correcting for multiple testing (*p*_*FDR*_<.05).

### Genetic liability to cannabis use predicts mental health/risk behavior outcomes and vice versa

Polygenic scores (PGS) were created in an independent sample (N_max_=32,033) from the Swedish Twin Register to assess if genetic liability to ever-use (trans-ancestry and European-only), and the *All-frequency* and *All-cannabis* factors were associated with 24 substance use and mental health outcomes (full results in <u>Table S49</u>). The PGSs for ever-use were significantly associated with ever-use of cannabis, panic disorder, bipolar disorder, and regular smoking, with the latter three only showing significant associations for the trans-ancestry PGS, and with modest effect sizes. The *All-frequency* PGS was associated with phobia, ever-use of cannabis, ecstasy, and stimulants, regular smoking, and alcohol abuse (Figure 4). The *All-cannabis* PGS explained more substantial amounts of variance, with significant associations for 19 of the 24 traits, whereby higher PGSs predicted more substance use and worse mental health. The strongest associations for the *All-cannabis* PGS were observed with schizophrenia (R^2^=4.16%, 95% confidence interval CI=0.28-12.49%), ever-use of stimulants (R^2^=3.86%, CI=2.50-5.60%), cannabis (R^2^=3.09%, CI=2.19-4.12%), and ecstasy (R^2^=2.64%, CI=1.12-4.73%).

To assess the direction of possible causality underlying observed associations with mental health traits, we conducted instrumental variable analyses using MR (Figure 6, <u>Table S51</u>). We compared associations with ever-use and a combined *All-frequency*/CUD (instruments from the combined *All-frequency* and CUD GWAS to obtain sufficient instrumental variables). We found strong evidence (i.e., effects consistent across sensitivity analyses and surviving multiple testing correction) for a causal effect of cannabis ever-use on bipolar disorder risk and for an effect of *All-frequency*/CUD on schizophrenia, bipolar disorder, anxiety, and insomnia. There was suggestive evidence for causal effects of ever-use on anorexia nervosa and of *All-frequency*/CUD on PTSD risk and lower educational attainment. In the other direction, we found strong evidence for an effect of genetic risk for ADHD, MDD, PTSD, and higher educational attainment on ever-use, and of ADHD, MDD, bipolar disorder, schizophrenia, PTSD and lower educational attainment on *All-frequency*/CUD. There was suggestive evidence for effects of schizophrenia and bipolar disorder on ever-use, and anxiety on *All-frequency*/CUD. For well-being, obsessive-compulsive disorder, IQ, and loneliness we did not detect any consistent effects.

**Figure 6.**
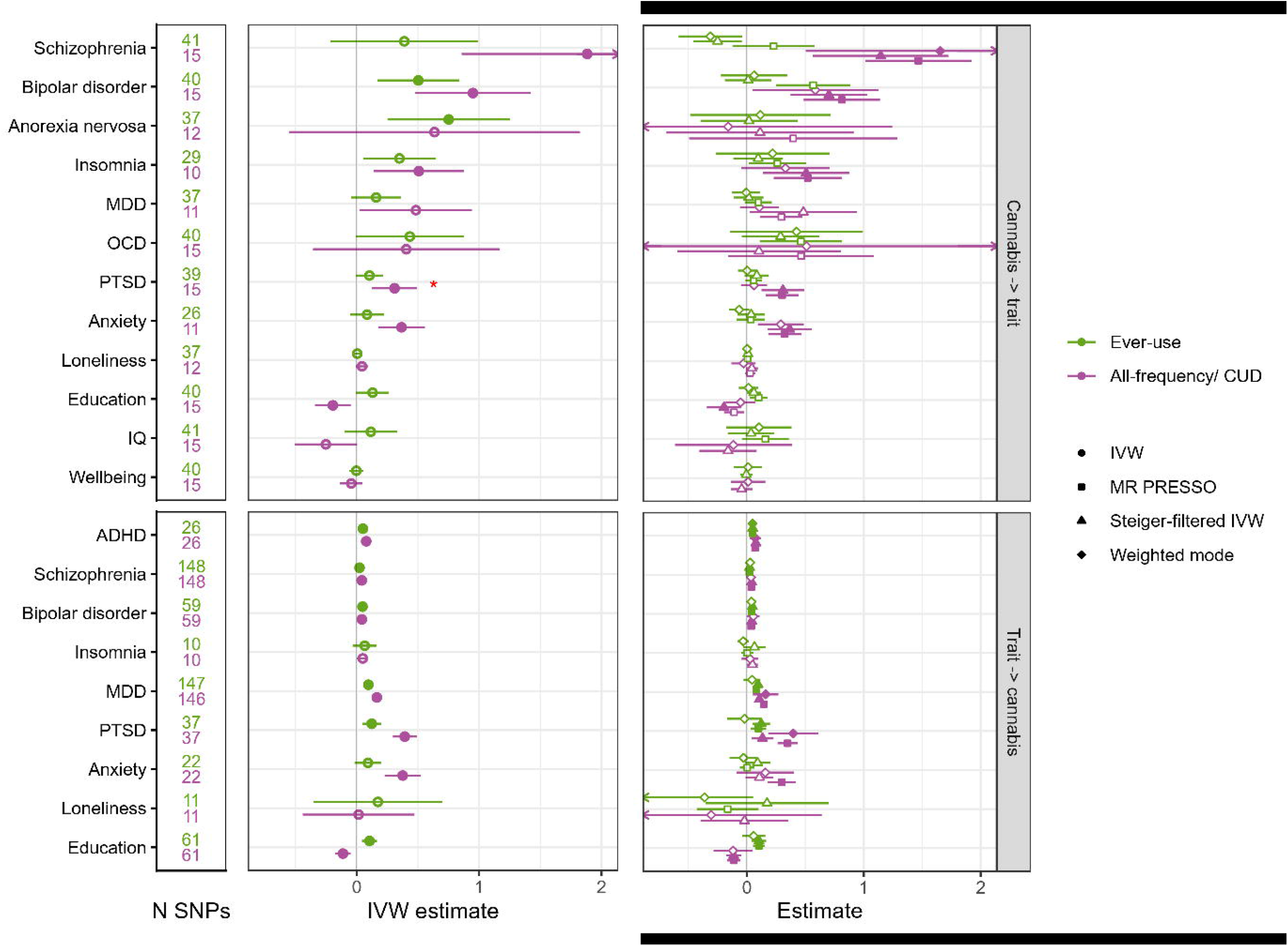
Mendelian randomization (MR) analysis results with the top panels showing effects of cannabis traits on the selected outcomes (<u>Table S52</u>), and the bottom panels showing the effect in the other direction (for the exposures with a sufficient number of instrument SNPs). The left panel indicates the number of instrument SNPs per analysis (N SNPs). In the middle panel the Inverse Variance Weighted (IVW) effect size is given with its CI. The right panel shows estimates for selected sensitivity analyses (all other results are given in <u>Table S51</u>). Note that estimates cannot be compared directly because the scale at which the outcomes were measured varied (e.g., binary vs. continuous; standardized vs. non-standardized) and because IVW estimates can be biased upwards when instruments have small effect sizes in the GWAS. Closed circles denote estimates that were significant at *p*_*FDR*_<0.05. * For this analysis, the MR Egger intercept was significant (*p*<0.05), indicating significant pleiotropy and limited interpretability. *MDD = major depressive disorder; OCD = obsessive-compulsive disorder; PTSD = posttraumatic stress disorder; IQ = intelligence quotient; ADHD = attention deficit/hyperactivity disorder; IVW = Inverse-variance weighted*

## Discussion

We conducted the largest genome-wide association study of cannabis ever-use traits to date, including information from over 736,000 individuals across ancestry groups (24% American, South Asian, East Asian, African, and other ancestries). We identified 54 independent loci for ever-use and 6 loci for frequency-related phenotypes. Moving beyond single phenotypes, we leveraged the availability of multiple measures of use to model the genetic architecture cannabis involvement in more depth. A latent *All-frequency* factor captured shared liability across lifetime, maximum, and recent use, and, together with CUD, formed an *All-cannabis* factor that indexes general liability across stages of use. Our multivariate approaches increased discovery and provided a more integrative view of the genetic architecture than any single phenotype alone. Across the ever-use, *all-frequency*, and *All-cannabis* GWASs, we identified 75 novel loci that were not implicated in previous GWASs of ever-use^5^ or CUD^6^ and implicated 446 different genes. We demonstrate both shared and trait-specific associations between psychiatric disorders and stages of cannabis involvement.

The SNP heritability of ever-use (5.4-6.5%) was lower than what we found in our previous International Cannabis Consortium GWAS^5^ (11%) but higher than the most recent estimate of 3.7%^9^. The reduction potentially reflects increased phenotype heterogeneity due to the inclusion of additional cohorts with increased variability in assessment instruments, age, cultural context, and legal status of cannabis use across samples. The trans-ancestry GWAS increased power to detect significant associations, but its heritability was lower than the European-only GWAS. This could be due to the use of reference data that was not ancestry-matched to estimate LD, but it is also plausible that substance use traits are environmentally contingent and context-dependent, with greater heterogeneity across birth cohorts, legal status, cultural norms, and ancestral backgrounds, thereby diluting cross-ancestry genetic signals and limiting gains in power.

We identified 446 unique significant genes (tier 3) across all cannabis use traits, most of which were associated with the well-powered ever-use trait and the latent *All-cannabis* factor. CADM2 was the most strongly implicated gene among the high-confidence candidates. Multiple previous GWASs of cannabis ever-use have also identified CADM2 as the top signal^5,8,9^. Variants mapped to this gene have been linked to risk⍰ taking, impulsivity, smoking initiation, and alcohol consumption^14-17^, providing additional support for *CADM2* influencing a general risk ⍰ taking personality trait that predisposes to substance use. We also identified several other high confidence genes within the 16p11.2 locus, including *TUFM, SULT1A2, ATP2A1, SULT1A1* and *NFATC2IP*. Genetic variation within this locus has not previously been linked to substance use behavior, but has been implicated in several neurodevelopmental disorders, including autism spectrum disorder, intellectual disability, epilepsy, and schizophrenia^18^.

Our gene set enrichment analyses implicated biological categories related to synaptic signaling (e.g., negative regulation of adenylate cyclase GPCR signaling), dendritic architecture, and neurodevelopment. These enriched biological categories broadly mapped onto cell types with significantly enriched heritability for cannabis use traits. For example, genetic signals for ever-use and *All-cannabis* were enriched in excitatory neurons in the amygdala and multiple hippocampal cell types, as well as superficial and deep cortical pyramidal neurons (e.g., deep layer intratelencephalic neurons), reflecting the widespread synaptic effects of cannabinoid drugs^19^. Gene set enrichment related to dendritic architecture is supported by experimental evidence that suggests chronic cannabinoid exposure remodels dendritic trees and spine density in prefrontal and accumbal neurons^20^. Adolescent THC exposure also prunes dendritic spines and alters gene networks in layer III pyramidal neurons^21^, supporting the enrichment seen in upper ⍰ layer intratelencephalic neurons. We also found enrichment of genetic signals for ever-use and *All-cannabis* in developmental pathways such as brain morphogenesis and somatic stem cell maintenance, which may reflect the role of endocannabinoid signaling in neurogenesis and axonal guidance^22^. This aligns with heritability enrichment in dentate gyrus and CA4 neurons, where cannabinoids have been found to promote neurogenesis and structural changes^23^.

By mapping shared and distinct components in the genetic liability to cannabis use and related traits, we can advance insight into biological mechanisms linking phenotypes and weigh in on the ongoing endeavor at identifying domain-general and domain-specific vulnerability (e.g., ^13,24,25^). We know from previous studies that ever-use and CUD have partly distinct genetic architectures^8,26^; here, we extend previous findings by adding new GWAS results for frequency of use, allowing for a more comprehensive genetic exploration across the different stages of cannabis use. We applied a wide range of methods (LDSC, Genomic SEM, GNA, MiXer, PGS) and made several interesting observations which will be summarized below.

First, our results demonstrated moderate to high genetic correlations (ranging from 0.60-0.76) across the three measures of use (ever-use, frequency of use, and CUD), which were adequately captured by a higher-order common-factor model representing a latent *All-cannabis* liability. This is consistent with twin studies of substance use progression, which have shown substantial overlap in the genetic influences underlying initiation, escalation, and abuse^2,27,28^.

Second, a comparison of the genetic architectures of the three traits revealed that SNP-based heritabilities ranged from 4.1% for *All-frequency* to 6.5% for ever-use and 6.7% for CUD^6^. MiXeR analyses showed that frequency of use is the most polygenic trait (i.e., is influenced by the largest number of causal variants). Nearly all of the causal variants associated with frequency of use (∼11,000) were shared with ever-use (∼8,000), and a substantial proportion were also shared with CUD (∼7,000), whereas direct overlap between ever-use and CUD was much lower. Of interest, conditional genetic modelling using GNA^29^ showed that the genetic correlation between ever-use and CUD was fully mediated by frequency of use. Taken together, our findings suggest that frequency of cannabis use captures much of the shared genetic liability linking ever-use and CUD. Whereas ever-use shares genetic liability with both traits, its association with CUD appears largely indirect and is consistent with the idea that use frequency is on the causal pathway from ever-use to CUD^30^. This may reflect that ever-use is mainly driven by novelty seeking and other personality traits, while frequency is more related to addiction proneness.

Third, we examined genetic correlations with external traits. All cannabis traits showed positive genetic correlations with a broad range of mental health and substance use traits, with associations generally strongest for CUD. The patterns of genetic correlation for *All-frequency* more closely resembled those for ever-use than for CUD, although a dose-response pattern was observed for some traits, such as age at smoking initiation and smoking cessation. Overall, these results indicate increasing genetic correlations with psychopathology and substance-related liability as cannabis involvement increases. Notably, while CUD was negatively genetically associated with several cognitive and socioeconomic outcomes (consistent with an average decline of ∼2 IQ points following frequent or dependent cannabis use in youth^31^) ever-use showed modest positive correlations with favorable outcomes, whereas *All-frequency* showed very weak associations. Moreover, all of the three cannabis traits showed negative correlations with conscientiousness and positive correlations with openness, but the associations were stronger for ever-use. Overall, these results indicate that recreational cannabis use and dependence reflect overlapping but partly distinct genetic profiles, although differences in ascertainment and participation across samples may also have contributed to the observed patterns. While ever-use is characterized by stronger associations with certain social or personality traits (e.g., risk-taking, externalizing behavior, and openness (in line with^13,17^)), CUD is more strongly associated with substance use disorders and internalizing mental health conditions such as anxiety, MDD, and PTSD.

Fourth, we calculated PGS in an independent Swedish target sample. The *All-cannabis* PGS was associated with increased risk of substance use and mental health problems, while ever-use was significantly associated with only a few traits and explained little variance. *All-frequency* significantly predicted some of the outcomes (e.g., ever-use of ecstasy and stimulants), but the associations were attenuated compared to those observed for *All-cannabis*. This suggests that associations with adverse outcomes are driven by more serious forms of cannabis use, although the observation that *All-cannabis* was the strongest also likely reflects that its source GWAS was the most powerful. Interestingly, the ever-use PGS based on the trans-ancestry GWAS had better explanatory power than the one based on the European-only GWAS, even though SNP-heritability and number of risk loci were similar in these GWASs, and even though the target sample was European-only. The trans-ancestry GWAS likely provided more accurate SNP effect estimates by leveraging differences in linkage disequilibrium (LD) structure across populations, which can improve tagging of causal variants and the calibration of effect sizes^32,33^.

The MR analyses revealed strong evidence for bidirectional associations between cannabis *All-frequency*/*CUD* and both bipolar disorder and schizophrenia. We also observed unidirectional causal associations from ever-use to bipolar disorder and from *All-frequency*/*CUD* to anxiety and insomnia. In the other direction, ADHD, MDD, PTSD and higher education predicted ever-use, while ADHD, MDD, PTSD, and lower education predicted *All-frequency*/*CUD*. The reverse direction for the association with education may reflect a similar pattern as previously observed for alcohol use, where ‘normative’ use is positively associated with socioeconomic status, while abuse has a negative relationship^34^. Notably, *All-frequency*/*CUD*, but not ever-use, was causally associated with increased schizophrenia risk, suggesting a potential dose-response relationship. The causal associations from mental health disorder to cannabis use are consistent with cannabis being used as a form of self-medication to alleviate psychiatric and/or somatic symptoms^35^. As a caveat, these findings could be influenced by weak instrument bias in the cannabis GWAS, given the typically weaker genetic instruments for cannabis traits relative to other psychiatric disorders. Note that when using MR on complex (behavioral) traits like cannabis use, especially using an explorative approach as we did here, the results are not conclusive and require triangulation using other methods and data^36^. An important limitation is that we could not construct a strong genetic instrument for cannabis frequency alone. Therefore, we combined the frequency GWAS with CUD to obtain a sufficient number of genome-wide significant variants. This composite phenotype complicates interpretation, as it blends traits.

More generally, this study should be interpreted in the context of some important limitations. First, the cannabis use traits were based on self-report, which can result in recall bias and potential under-reporting stemming from social desirability or legal concerns. Also, the use of the binary ever-use phenotype may capture heterogeneous patterns of use, which could dilute associations related to experimental single use, recreational use, frequent or disordered use. The cannabis frequency measures differed across cohorts. We grouped them into three harmonized categories (lifetime, maximum, and recent frequency), which could be captured in a single common factor with good model fit. Nevertheless, substantial heterogeneity remained, both within and between the different frequency indices. This additional measurement error likely attenuated the genetic signal and may have contributed to lower SNP heritability estimates. Furthermore, while 24% of the sample was of non-European ancestry, which is a significant advance over previous GWASs, the limited statistical power in this subgroup, as well as the reliance of most downstream methods on single-ancestry reference data meant that many of the post-GWAS analyses were based on the European-ancestry results. Finally, despite the large overall sample size, the cannabis frequency phenotypes were available in a substantially smaller subset, reducing power for those analyses and for the downstream applications involving polygenic scores and Mendelian randomization.

In conclusion, this study represents the largest GWAS meta-analysis of cannabis use traits to date, encompassing a sample of over 736,000 individuals. It identified 47 genome-wide significant SNPs for ever-use and 6 for frequency of use. By triangulating evidence across four complementary analytic approaches, we identified a set of high-confidence genes likely involved in cannabis use. We further showed that ever-use, frequency of use, and CUD have overlapping but partially distinct genetic architectures, with frequency of use capturing most of the shared genetic liability between initiation and CUD. Also, ever-use was genetically linked to increased risk-taking and higher socioeconomic status, whereas measures of cannabis use frequency and CUD had stronger associations with psychiatric disorders and adverse outcomes. MR showed evidence of bidirectional causal associations between ever-use and bipolar disorder, and between heavier cannabis involvement and schizophrenia. These findings are in line with a dose-response relationship with schizophrenia risk and are consistent with self-medication. MR also identified unidirectional effects from heavier cannabis use to PTSD, and from ADHD to cannabis use. Combined, these results advance our understanding of the genetic architecture of cannabis use traits, as well as relationships with other psychiatric disorders and complex traits.

## Online methods

### Cohorts and traits

For **ever-use**, cases were individuals who had used cannabis at least once in their lifetime, controls had never used cannabis. For frequency of use we analyzed three different measures (in lifetime users only): 1) **lifetime frequency**; total number of times used during lifetime), 2) **maximum frequency**; frequency of use during period that the individual used most often, and 3) **recent frequency**; frequency of use in the past year/month(s). There were variations on how exactly these traits were measured in the cohorts (<u>Table S53</u>), but all used survey items with response categories.

We used data from 34 cohorts with a total sample size (with any of the cannabis traits) of N=736,322. For ever-use (yes/no), the European sample size was N=556,258 (with 273,372 cases). The largest contributing cohorts were from the 23andMe Research Institute, All of Us, and UK-Biobank. For the trans-ancestry analyses, we also included N=64,570 American, N=71,898 African, N=8,546 East-Asian, N=3,623 South-Asian, and N=29,730 other or admixed descent individuals from the HELIUS and All of Us cohorts. Here, genetic ancestry was inferred from individual-level genetic data by comparing it to reference data and defined five broad continental categories, one category defined as “other” that includes individuals belonging to ancestry category with small sample size, and one category defined as “admixed” that includes individuals with genetic admixture. We acknowledge that genetic ancestry exists on a continuum and that any form of categorization is suboptimal and an oversimplification, which may reinforce typological thinking about human differences^37^. We refer to these groups concurrently with the acronym *AmSEAfO* (Am=American, S=South-Asian, E=East-Asian, Af=African, O=other/admixed), and to analyses combining European with *AmSEAfO* samples as *trans-ancestry*. European sample sizes for lifetime, maximum, and recent frequency were N=73,743, N=136,682, and N=85,147, respectively. For recent frequency, data were also available for individuals with an *AmSEAfO* ancestry in the All of Us cohort (N=42,163). Descriptions for all cohorts are included in <u>Methods S1</u>.

### GWAS protocol

All cohorts followed a standardized analysis protocol. Quality-control involved excluding SNPs with a call rate <95%, minor allele frequency (MAF) <1%, Hardy-Weinberg disequilibrium p<1E-6, and individuals with chromosomal abnormalities, excessive autosomal homozygosity or heterozygosity, and sex abnormalities. SNPs were imputed to the Haplotype Reference Consortium panel data. GWA tests were run while controlling for age, sex, birth cohort (dummy-coded in 20-year cohorts), principal components (PCs) capturing population stratification effects (number of components as appropriate for the study population), and if necessary, study-specific covariates, such as array, batch, or site-effects (see <u>Table S54</u> for details per cohort). GWASs were run separately per ancestry group.

### GWAS meta-analyses

For the European ancestry GWASs, we conducted four meta-analyses (for ever-use, lifetime frequency, maximum frequency, and recent frequency) using METAL^38^. For the ever-use phenotype, we combined the GWAS effect sizes weighted by the inverse standard errors. For the frequency traits, we weighted by sample size, because there were measurement differences in the cohorts. The X chromosome was analyzed separately, because it was available for only some cohorts and sample size was sufficient only for the ever-use phenotype.

For ever-use, data were available from individuals of European and *AmSEAfO* ancestries in the HELIUS and All of Us cohorts. Trans-ancestry GWAS was conducted using GENESIS^39^, which models population structure, relatedness, and admixture without relying on manual ancestry classification. The *AmSEAfO* GWAS results were meta-analyzed with the European meta-analysis using METAL.

For the meta-analyses results, LDscore regression (LDSR)^40^ was used to assess how much of the variance in the phenotype was explained by the SNP-effects captured in the analysis (SNP-based heritability, h^2^_SNP_). For the trans-ancestry meta-analysis we used European LD-reference data, as it was the largest group in the analysis. As a sanity check, we also estimated h^2^_SNP_ based on LD-reference data from the *AmSEAfO* part of the All of Us sample. Using *AmSEAfO* reference data, h^2^_SNP_ estimates were somewhat lower (-1.4% for ever-use and -0.2% for recent frequency), which is likely due to stratification effects in this mixed dataset.

To assess if the genetic liability to cannabis use was similar in different ancestries, we used Popcorn^41^ to estimate genetic correlations for the ever-use and recent frequency GWASs. Because the separate non-European ancestry cohorts were small, we estimated the correlation between European and combined *AmSEAfO* results. We used reference data from 1000 Genomes, with the combined ancestry panel (roughly equal samples of European, African, East Asian, and admixed American ancestry) as reference for the *AmSEAfO* cohort. This ancestry composition is not the same as in our *AmSEAfO* analysis, but is a better representation of it than European-only reference data would be. Because the *AmSEAfO* results for ever-use had a non-significant heritability when including the HELIUS cohort, we used the results based on the All of Us cohort only to estimate this genetic correlation.

Note that for all other follow-up analyses, we were restricted to the European-ancestry results due to sparsity of trans-ancestry databases, reference data (with the same ancestry composition as our GWASs), and methods equipped to deal with trans-ancestry data.

### Modelling genetic relationships between cannabis use traits

Continuing with the European ancestry results, we explored how different cannabis traits are expressions of the same underlying genetic liability. We visualized the genetic relationships between the three frequency traits based on transformations from GDIS^42^. Specifically, we used LDSR to estimate heritability of the European-ancestry frequency GWASs and their genetic correlations, and then visualized this information using the square root of the heritabilities to determine the length of the vectors and the inverse cosine of the genetic correlations to determine their angles. The vectors were then projected into 3D Euclidean space based on these parameters.

Following the results from GDIS and because we wanted to contrast ever-use to frequency in the follow-up analyses, we proceeded to conduct a multivariate GWAS of the three frequency traits. We performed meta-analysis in Genomic Structural Equation Modeling (Genomic SEM^43^), which also uses the genetic covariance matrix but relies on a SEM framework. We refer to the results of this GWAS as ***All-frequency***.

We then added summary statistics from an existing GWAS on CUD^6^ (using the European ancestry sample, N=886,025, N-cases=42,281) to investigate common and distinct liability to different stages of use. First, we estimated the bivariate genetic correlations between all cannabis use traits using LDSR. Second, we used MiXeR^44^ (v1.3) to quantify polygenicity (the total number of trait-influencing genetic variants) and polygenic overlap (the total number of variants that influence both of a pair of traits) among the cannabis ever-use, cannabis frequency, and CUD. MiXeR fits a Gaussian mixture model assuming that common genetic effects on a trait are a mixture of causal variants and noncausal variants. Polygenicity is reported as the number of causal variants that explain 90% of SNP heritability of the trait (to avoid extrapolating model parameters into the area of infinitesimally small effects).

Finally, we parsed the common and independent genetic signal for ever-use, *All-frequency* (the GWAS combining the three frequency traits), and CUD using Genomic Network Analysis (GNA)^29^. This method models the conditional association between all trait pairs, relying on the genetic correlation matrix estimated with LDSR. Third, we tested how well the data fit a common factor model constituted by the different cannabis traits. We call this common factor of ever-use, *All-frequency*, and CUD ***All-cannabis*** and perform a GWAS on this factor in Genomic SEM. Finally, we repeated the common factor GWAS leaving out ever-use for the Mendelian randomization analysis (discussed below). This GWAS we call ***Frequency/CUD***.

### Gene prioritization

We triangulate across four different methods to identify high-confidence genes that are likely involved in cannabis use, focusing on the ever-use, *All-frequency*, and *All-cannabis*.

#### MAGMA

MAGMA (Multi-marker Analysis of GenoMic Annotation)^45^ was used to identify risk genes near genome-wide significant loci for the three cannabis traits. This approach assigns variants to genes based on their physical proximity to a defined set of genic coordinates. We mapped variants to protein coding genes obtained from the MAGMA website (https://cncr.nl/research/magma/). Genic coordinates extended 5 kilobases (kb) upstream and 1.5 kb downstream to capture regulatory variation. After mapping variants to genes, gene-based P-values were calculated using the SNP-wise mean model. We used samples from the 1000 Genomes Project (Phase 3) as the LD reference panel.

#### mBAT-combo

We performed a gene-based analysis utilizing the multivariate Set-Based Association Test (mBAT-combo^46^) within GCTA version 1.94.1. This method enhances the detection power for multi-SNP associations, especially in the presence of masking effects, by combining mBAT and fastBAT test statistics through a Cauchy combination approach. This allows for the integration of different test statistics without prior knowledge of their correlation structure, thereby maximizing overall power regardless of masking effects at specific loci. We used the European subsample (N=503) from Phase 3 of the 1000 Genomes Project as the LD reference panel, applying the fastBAT default LD cut-off of 0.9. A gene list of 19,899 protein-coding genes was utilized to map base pair positions according to genome build hg19.

#### Transcriptome-wide association study

We performed a transcriptome-wide association study (TWAS) using the FUSION method, which integrates GWAS summary statistics with gene expression prediction models to identify associations between gene expression and cannabis use. We utilized brain gene expression weights from the PsychENCODE project, which includes expression data from the dorsolateral prefrontal cortex. We incorporated LD information from the 1000 Genomes Project Phase 3 to estimate the genetic component of gene expression. This genetic component was then tested for association with each cannabis use phenotype using GWAS summary data. To correct for multiple testing, we applied the Bonferroni correction (i.e., 0.05/number of tests performed). We used the COLOC R package^47,48^, implemented using FUSION, to identify TWAS associations (i.e., trait-gene expression associations) that are likely to share a causal SNP. Colocalisation calculates posterior probabilities (PP) to assess whether individual lead SNPs within a significant TWAS locus are: (i) independent signals (PP3) (e.g., two causal SNPs in LD, one affecting transcription and the other affecting cannabis use), or (ii) shared signals (PP4) (e.g., a single causal SNP affecting both transcription and cannabis use).

#### Summary-based Mendelian Randomization

We used Summary data-based Mendelian Randomization (SMR) to integrate GWAS and expression quantitative trait loci (eQTL) data from human brain and blood samples. This method leverages summary-level data in an MR statistical framework to test for pleiotropic associations between gene expression and GWAS data. We used whole blood expression data from the eQTLGen consortium^49^ and brain expression data from the MetaBrain resource^50^. To assess the presence of linkage or pleiotropy, we performed the Heterogeneity in Dependent Instruments (HEIDI) test alongside SMR, evaluating effect size heterogeneity between the GWAS and eQTL summary statistics. The HEIDI test is designed to distinguish between pleiotropy and linkage by assessing heterogeneity in the association signals. A HEIDI P-value threshold of 0.05 was used to determine the likelihood of linkage influencing the observed association.

#### High confidence gene sets

To identify high-confidence gene-level associations for each cannabis phenotype, we integrated results across different gene-based tests. For each phenotype, we first compiled association statistics from MAGMA, mBAT-combo, TWAS with GTEx whole blood, TWAS with PsychENCODE/PEC transcriptomes, SMR with eQTLGen whole blood, and SMR with BrainMeta cortical eQTL resources. Gene-based test-specific significance thresholds were applied using Bonferroni correction (*p*_*Bonferroni*_<0.05). For expression-based tests, we additionally required strong evidence that the association reflected a shared causal variant: TWAS signals were retained only when supported by colocalisation (PP4≥0.8), and SMR associations were required to pass the HEIDI test (*p*_*HEIDI*_>0.01) to exclude heterogeneity due to linkage.

For each gene we then defined independent lines of evidence across three domains: (i) proximity gene-based GWAS evidence (significant association in MAGMA or mBAT-combo); (ii) functional brain-derived expression evidence (significant TWAS–PEC with colocalization and/or significant SMR– BrainMeta with HEIDI support); and (iii) functional blood-derived expression evidence (significant TWAS–GTEx with colocalization and/or SMR–eQTLGen with HEIDI support). A gene was classified as high-confidence for a given phenotype when it was supported by at least one proximity gene-based GWAS test and at least one functional expression-based result with colocalization/HEIDI support from any tissue. Genes supported only by gene-based tests or by at least 2 functional expression-based tests were retained as secondary “Tier 2” candidates. The remaining genes with Bonferroni-significant evidence in at least one gene-based test that did not meet the criteria for Tier 1 or Tier 2 were classified as Tier 3 candidates. High-confidence sets were generated independently for each cannabis phenotype.

### Enrichment and functional annotation

To aid interpretation of the GWAS findings, we performed enrichment analyses assessing functional implications and plausible biological pathways for the loci involved in cannabis use behavior.

#### Partitioned Heritability

We used stratified LDscore regression^51^ to assess if the heritability of ever-use, *All-frequency*, and *All-cannabis* was enriched in 30 human brain cell superclusters as mapped in the Adult Human Brain Atlas^52^. We compared brain cell types involved in ever-use and *All-frequency*, and included the *All-cannabis* trait to increase power and assess which cell types are generally involved in cannabis use.

#### Gene set enrichment analysis with GSA-MiXeR

Gene set analysis was conducted using GSA-MiXeR to quantify the enrichment of genetic signals across ever-use, *All-frequency*, and *All-cannabis* use traits within gene ontology (GO) categories. We applied default parameters as described by Frei et al.^53^. GSA-MiXeR estimates the s-fold enrichment of heritability in pre-defined gene sets through maximum likelihood optimization, while accounting for LD, MAF, and polygenicity. To present the results, we first identified significant (*p*_*FDR*_<0.05) gene sets using competitive gene set enrichment with MAGMA. Gene ontology sets with MAGMA *p*_*FDR*_<0.05 were then ordered based on their fold enrichment of genetic signal for each cannabis use phenotype from GSA-MiXeR.

### Look-up strategy

The top SNP in each genomic risk locus for the different GWASs was queried in GWASAtlas (https://atlas.ctglab.nl/PheWAS) to identify associations with other traits in previously published GWASs through a phenome-wide association study (PheWAS). The PheWAS table for each SNP was downloaded and filtered to only include genome-wide significant SNPs.

In addition, the genomic risk loci were compared to the previous GWASs of ever-use^5^ (note that the summary statistics of the newest ever-use GWAS^9^ were not yet available at the time of study) and CUD^6^ to identify novel loci. We used a pipeline which compares summary statistics files to identify which loci were significant in both, one, or neither of the GWAS summary statistics files and provides the novel loci as output. Independent genomic risk loci were identified using clumping as implemented in the FUMA pipeline^54^ with R^2^<0.1 and distance>250kb and 1000 Genomes reference data (European sub sample).

Finally, based on the gene prioritization results, high-confidence genes were queried. We conducted a search of the top 20 tier 1 and tier 2 genes for ever-use and *All-frequency* (Figure 3B) using GWAS Catalog. Each Gene Symbol was input into the search interface, with a description of the gene from ENSEMBL, location of the gene on the reference genome and cytogenetic band corresponding to genomic location taken from the ‘Gene information’ panel on the individual gene pages. Information on all previously reported associations with the gene recorded in the GWAS Catalog was extracted. We documented these previously reported associations and derived the number of associated reported traits.

### Genetic correlations with other traits

Using LDSR, we explored genetic correlations between the cannabis traits with a set of 44 GWAS summary statistics broadly reflecting socioeconomic status, cognition, mental health, personality, risk taking, and substance use traits. The GWAS sources are listed in <u>Table S48</u>. We again compared ever-use to *All-frequency* and CUD.

### Polygenic score prediction

In an independent sample we created polygenic scores (PGS) to test if genetic predisposition to cannabis use was associated with observed mental health and substance use. We created scores for ever-use, *frequency*, and *All-cannabis* using LDpred2 (taking into account the LD structure in the population using 1000 Genomes reference data). For ever-use, we created a PGS based on the trans-ancestry GWAS as well as the European-only GWAS to assess if including trans-ancestry information improved explanatory power. Because LD structure cannot be modelled simultaneously across ancestries, we used European LD reference data for the trans-ancestry PGS.

The target sample included 32,033 individuals from the Swedish Twin Register (born 1959-1985, M=1971, SD=7.8, 59% female). We tested the effect of cannabis use PGS on 10 key mental health and 14 cannabis and other substance use traits (measures and sample sizes <u>Table S50</u>). To estimate the effect size of the PGS, we compared models with and without the PGS to derive the difference in explained variance (R^2^-change). Uncertainty in R^2^-change was obtained by nonparametric bootstrap with 2,000 resamples. For each PGS-outcome pair we reported the bootstrap 95% bias-corrected and accelerated (BCa) confidence interval (CI), which accommodates the skewed, bounded distribution of R^2^-change. When the BCa interval could not be computed (e.g., nonconvergent jackknife replicates), we reported the 95% percentile bootstrap interval instead. Significance of R^2^-change>0 was evaluated from the same bootstrap distribution (one-sided). We corrected for multiple testing using false-discovery rate p-values (correcting for the number of tested outcomes).

### Mendelian randomization

In Mendelian Randomization (MR), SNPs that are genome-wide significantly associated with a trait are used as instrumental variables for that trait (the ‘exposure’). Because SNPs are randomly segregated at conception and cannot be influenced by confounders, their effects can be interpreted as causal (given that certain assumptions are met). We tested putatively bidirectional causal relationships between cannabis ever-use and a set of 14 selected traits (<u>Table S52</u>). *All-frequency* could not be used as an exposure because it identified fewer than 10 instrument SNPs, limiting power for MR analysis. Because we wanted to compare with ever-use, we chose not to use the *All-cannabis* common factor GWAS. This trait combined conditionally dependent traits (risk variants for frequency and CUD might not have the same meaning for users versus non-users), and was therefore replaced by a common factor GWAS combining *All-frequency* and CUD for this analysis.

Traits were selected based on a) previous literature suggesting a plausible causal relationship with cannabis use (i.e., only plausibly temporally ordered relationships were tested, so not from cannabis use to childhood-onset disorders); b) the trait GWAS had a significant SNP based heritability with Z-score>7; c) there was a genetic correlation with at least one of the cannabis traits of *r*_*g*_>.20. The effect of the trait on cannabis outcomes was only tested if there were at least 10 genome-wide significant SNPs to be used as instrumental variables. Wherever possible, GWAS summary statistics excluding the UK-Biobank, the major source of overlap, were used. The only GWASs for which there were no summary statistics available leaving-out UK-Biobank were the ones for anxiety disorders and anorexia nervosa, but overlap was limited.

Some key MR assumptions can be tested and corrected for using sensitivity analyses. We used Steiger filtering, MR Egger regression, weighted median, and MR PRESSO^55^ to address pleiotropy and weak instrument bias. Steiger filtering excludes instrument SNPs that explain more variance in the outcome than in the exposure (at *p*<0.05) to reduce chances of reverse causality. MR Egger was used to assess horizontal pleiotropy (using MR Egger intercept *p*<0.05); in the case of a significant intercept, the IVW was not interpreted and the MR Egger slope was used instead. MR Egger regression corrects for detected pleiotropic effects and is reported alongside the estimates from the main tests to assess if it had consistent directions. The weighted median was reported as it is more robust to invalid or pleiotropic instruments. MR PRESSO detects pleiotropic outliers and recalculates the causal effect while excluding them one-by-one. Instrument strength was assessed using R^2^ and the F-statistic (values F>10 were considered sufficient), and heterogeneity in the effects across the SNP instruments was assessed using Cochran’s Q. For MR-Egger analyses, we calculated I^2^ to evaluate the reliability of SNP–exposure associations^56^. MR-Egger results were considered reliable when I^2^>0.9; Simulation Extrapolation (SIMEX) correction was applied when I^2^ values were between 0.6 and 0.9 to correct for bias. If values were below 0.6, MR Egger was not reported. The analyses were conducted using the TwoSampleMR^57^, MR PRESSO^55^, and simex packages in R.

We report the Inverse Variance Weighted (IVW) statistic as our main estimate of the effect. Its *p*-value was corrected using false discovery rate procedures^58^ within exposure, thus correcting for the number of independent outcomes. If the effect size direction was consistent with the IVW across the sensitivity analyses and mostly had p-values below 0.05, we interpreted that as strong evidence for a causal effect. If IVW was fdr-significant but results were not fully consistent across sensitivity analyses, we interpreted that as suggestive evidence.

## Supporting information

Supplementary Methods - Cohort descriptions

## Data availability

The full GWAS summary statistics excluding 23andMe as well as the top-10,000 SNPs including 23andMe will be shared via GWAScatalog upon publication. All code generated for the project will be shared via Figshare.

## Acknowledgements

We would like to thank the research participants and employees of 23andMe for making this work possible. The genome-wide association analysis on the UK Biobank dataset has been conducted using the UK Biobank resource under application number 40310. The HELIUS study is conducted by the Amsterdam University Medical Center, location AMC and the Public Health Service of Amsterdam. Both organizations provide core support for HELIUS. The HELIUS baseline measurement was additionally funded by the Dutch Heart Foundation, the Netherlands Organization for Health Research and Development (ZonMw), the European Union (FP-7), and the European Fund for the Integration of non-EU immigrants (EIF). The HELIUS follow-up measurement was additionally supported by the Netherlands Organization for Health Research and Development (ZonMw; 10430022010002), Novo Nordisk (18157/80927), the Swiss National Foundation (189235), University of Amsterdam (Research Priority Area 25-08-2020 “Personal Microbiome Health”), and the Dutch Kidney Foundation (Collaboration Grant 19OS004). The study reported here was partially supported by funding from the Dutch Cancer Society (VU2017-8288).

JAP was supported by the Amsterdam University Medical Center Postdoc Career Bridging grant (27527). ZFG was supported by National Health and Medical Research Council (NHMRC) EL1 Investigator Grant fellowship 2034743. ABT is supported by a PhD scholarship from Amsterdam University Medical Center. JSO is supported by an Australian NHMRC EL1 Investigator grant fellowship 2018420. HMS and REW are members of the MRC Integrative Epidemiology Unit at the University of Bristol which is supported by the Medical Research Council and the University of Bristol (MC_UU_00011/7). REW is also funded by a postdoctoral fellowship from the South-Eastern Norway Regional Health Authority (2020024). MPMB is supported by KNAW prize PAH/6635. IBH is supported by NHMRC Research Fellowship APP2016346: Right care, first time: delivering technology-enabled mental health care to young people at scale. SM is supported by National Health and Medical Research Council (NHMRC) L2 Investigator Grant fellowship 2034568. MN is supported by ERC Advanced Grant 2024. JLT is supported by a European Research Council (ERC) Starting grant (UNRAVEL-CAUSALITY, grant number 101076686). Views and opinions expressed are however those of the author only and do not necessarily reflect those of the European Union or the European Research Council. Neither the European Union nor the granting authority can be held responsible for them. EMD is supported by National Health and Medical Research Council (NHMRC) L1 Investigator Grant fellowship 2026364. K.J.H.V. is supported by the Foundation Volksbond Rotterdam. This work was supported by a 2024 BBRF Distinguished Investigator Grant (32124) from the Brain & Behavior Research Foundation and by the Netherlands Organization for Scientific Research (NWO) under the Open Competition grant (Grant No. 406.22.GO.043).

## Conflict of interest

SLE, PF, and SS are former employees of 23andMe. DD is a Co-founder and Chief Scientific Officer for Thrive Genetics, Inc. She is on the Advisory Board for the Seek Women’s Health Company and HumanUp. She has received royalties from Penguin Random House for her book, The Child Code: Understanding Your Child’s Unique Nature for Happier, More Effective Parenting. IH is a Professor of Psychiatry and the Co-Director of Health and Policy, Brain and Mind Centre, University of Sydney. He holds a 3.2% equity share in Innowell Pty Ltd that is focused on digital transformation of mental health services.

## Figure captions

**Figure 0.** Graphical abstract. Overview of analytical flow with figure thumbnails for the corresponding results. A more detailed overview of the included samples is given in <u>Table S1</u> and a summary of the GWAS analyses is given in <u>Table S2</u>.

## Notes

### Author Declarations

Each cohort whose data was used in this study has acquired relevant ethics permission covering this work. Ethics statements for all cohorts are included in Supplementary Methods S1.

