## Supplementary material for "Genetics of cannabis ever-use and frequency across ancestries implicate novel loci and brain-specific biology": Suplementary Figure 1-3

**Figure S1.** Manhattan plot of the p-values of each SNP association with ever-use in AmSEAfO ancestry only. If SNPs could be positionally mapped to their nearest genes, gene names are displayed. Top SNPs in each locus are annotated. The horizontal dashed lines indicate the genome-wide significance threshold of *p*<5e-8.

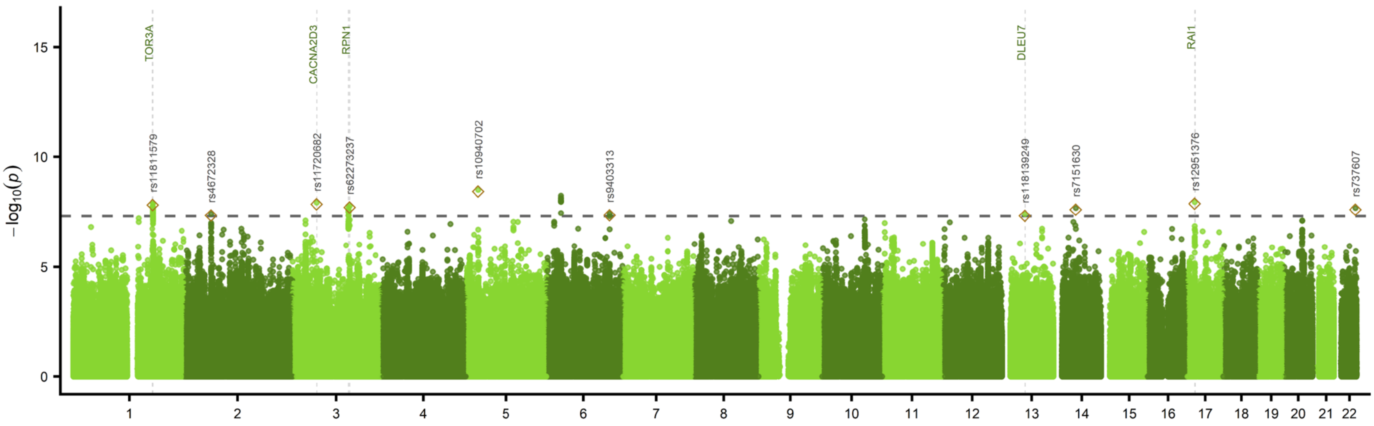

**Figure S2.** Manhattan plots of the p-values of each SNP association with frequency traits in European ancestry only. Top SNPs in each locus are annotated. The horizontal dashed lines indicate the genome-wide significance threshold of $p$<5e-8.

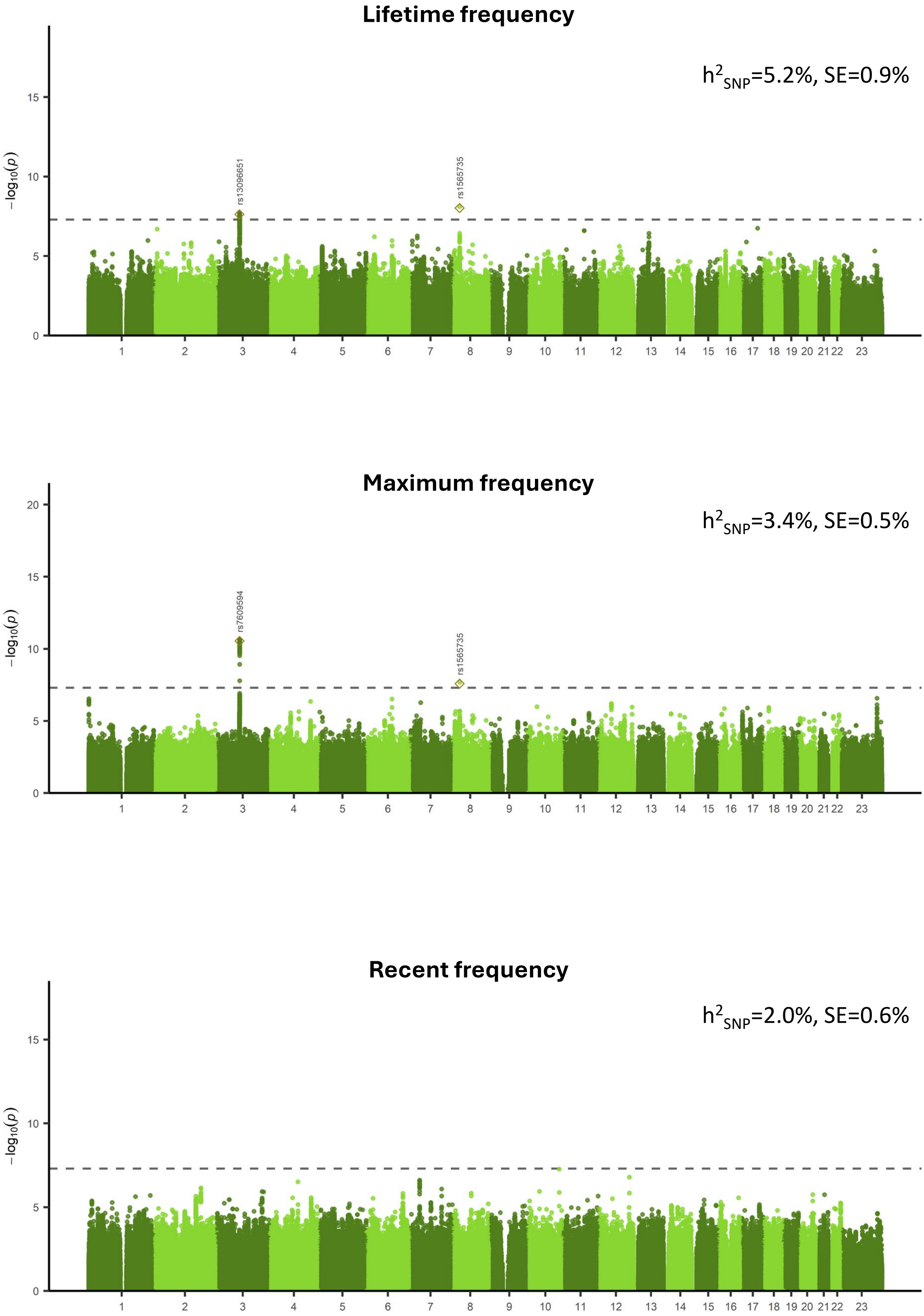

**Figure S3.** Manhattan plots of the p-values of each SNP association with recent frequency in *AmSEAfO* ancestry only. If SNPs could be positionally mapped to their nearest genes, gene names are displayed. Top SNPs in each locus are annotated. The horizontal dashed lines indicate the genome-wide significance threshold of $p$<5e-8.

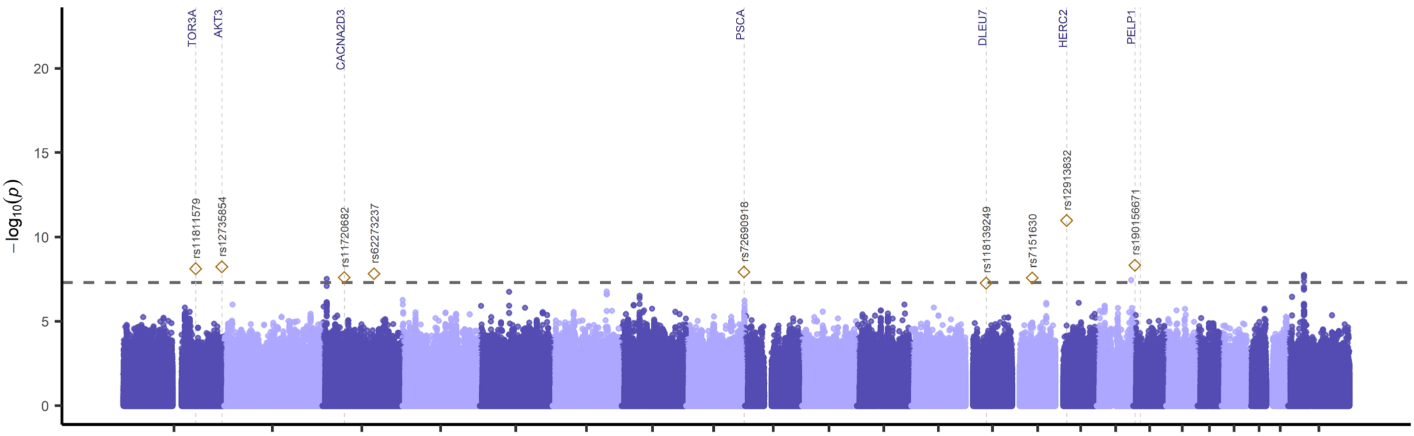
