## Supplementary Methods - Cohort descriptions for "Genetics of cannabis ever-use and frequency across ancestries implicate novel loci and brain-specific biology"

**Supplementary Methods S1. Cohort information.** Information on phenotype measures, genotyping, QC and imputation, and ethics and funding statements.

### 23andMe Research Institute

Cohort Description

23andMe Research Institute aims to make and support scientific discoveries about genetics and other factors behind diseases and traits. Over 80% of our members have consented to allow their de-identified research information (including genetic and self-reported data) to be used for research. Phenotypic data is self-reported by consented 23andMe research participants who have responded to web-based surveys or completed web-based assessments provided through 23andMe's Services.

Phenotype measure

As part of an online survey about decision making, we asked a series of questions about tobacco, alcohol, and other drug use, including cannabis. Ever use was assessed using the question, "Have you ever in your life used the following: Marijuana" (responses: Yes/No). Among ever users, recent use was assessed using the question, "How many days did you use the following substance within the last 30 days: Marijuana" (integer responses 0-30). Maximum use was similarly assessed among ever users, with the question, "How many days did you use the following substance during your heaviest 30 days: Marijuana" (integer responses 0-30).

Genotyping, QC, Imputation

Detailed methods for the recent use phenotype are described in Pasman et al 2018 (PMID 30150663), and for the ever use and maximum use phenotypes, in Thorpe et al 2025 (PMID 41077612). Briefly, DNA extraction and genotyping was performed on saliva samples by National Genetics Institute (NGI). Samples were genotyped on one of five Illumina SNP array designs with between 560,000 and 950,000 SNPs, and samples with call rate below 98.5% were rejected and repeated. For the recent-use GWAS, we imputed with a reference panel of 1000 Genomes Phase 3 and UK10K data. For the other phenotypes, we also imputed against a Haplotype Reference Consortium (HRC) panel, and merged results, with preference for the HRC result if a variant was present in both panels. Our GWAS quality control included flagging variants with low imputation quality (Rsq < 0.3), and variants with evidence of strong batch effects across genotyping platforms. We identified individuals with primarily European genetic ancestry (>97%) from aggregating local ancestry inference results. For a given phenotype, we identified related individuals using a segmental identity-by-descent (IBD) estimation algorithm. We used a greedy algorithm to exclude close relatives using a threshold of 700 cM IBD, roughly corresponding to the minimum expected sharing for first cousins,

Ethics Statement

Research participants provided informed consent and volunteered to participate in research online under a protocol approved by the external AAHRPP-accredited Institutional Review Board (IRB), Ethical & Independent (E&I) Review Services. As of 2022, E&I Review Services is part of Salus IRB (https://www.versiticlinicaltrials.org/salusirb)

Funding

AAP is supported by NIDA (P50DA037844 and P30DA060810). SSR is funded through the National Institute on Drug Abuse (NIDA DP1DA054394) and the Tobacco-Related Disease Research Program (T32IR5226)

Acknowledgments

We would like to thank the research participants and employees of 23andMe Research Institute for making this work possible. The following members of the 23andMe Research Team contributed to this study: Adam Auton, Alan Kwong, Anjali J. Shastri, Barry Hicks, Catherine H. Weldon, David A. Hinds, Emily DelloRusso, Emily M. Rios, Joyce Y. Tung, Kahsaia de Brito, Katelyn Kukar Bond, Keng-Han Lin, Matthew H. McIntyre, Matthew J. Kmiecik, Qiaojuan Jane Su, Robert K. Bell, Sayantan Das, Shubham Saini, Stella Aslibekyan, Vinh Tran, Wanwan Xu, Alisa P. Lehman, Noura S. Abul-Husn, R. Ryanne Wu, Rebecca M. K. Berns, Ruth I. Tennen, Stacey B. Detweiler, Aditya Ambati, Anna Guan, Bertram L. Koelsch, Chris German, Éadaoin Harney, Ethan M. Jewett, G. David Poznik, James R. Ashenhurst, Jingran Wen, Peter R. Wilton, Steven J. Micheletti, and William A. Freyman.

### All of Us (AoU) Research Program | PMID: 31412182

Cohort Description

The All of Us (AoU) Research Program aims to recruit a diverse cohort of at least one million individuals across the United States to advance biomedical research and enhance health outcomes [https://pmc.ncbi.nlm.nih.gov/articles/PMC8291101/]. The program enrolls participants aged 18 and older where they contribute data through surveys on lifestyle, demographics, and health history, as well as linked Electronic Health Records (EHR), physical measurements, and biospecimens.

Phenotype Measure

As part of lifestyle survey asking questions about use of tobacco, alcohol, and drugs, cannabis use data have been collected for the phenotypes below:

- Cannabis ever:

Cannabis ever use was assessed by asking: "In your LIFETIME, which of the following substances have you ever used?" and then providing "Marijuana (cannabis, pot, grass, hash, weed, etc.)" as one of the options.

- Cannabis recent frequency:

Cannabis recent frequency was assessed in ever users (selected marijuana in the previous question) by asking: "In the PAST THREE MONTHS, how often have you used marijuana (cannabis, pot, grass, hash, etc.)?"

Genotyping, QC, Imputation

We used the All of Us short read whole genome SNP & Indel smaller call set (ACAF v8). The genomic quality control is described at https://support.researchallofus.org/hc/en-us/articles/29390274413716-All-of-Us-Genomic-Quality-Report. In this study, GWAS was performed separately on participants with European genetic ancestry and non-European ancestry (that is, AFR, AMR, EAS, SAS, MID and OTH) using the AoU ancestry prediction. In both analyses, participants with discordant sex information, genotype missingness more than 5% and with at least one outlier QC metrics (provided by AoU) were excluded. Marker quality control included removing variants with more than 5% calls missingness. However, different minor allele frequency (MAF) cut-offs were applied for each analysis. MAF of < 1% for EUR subset versus < 0.5% for non-EUR subset.

Ethics Statement

Informed consent for all participants is conducted in person or through an eConsent platform that includes primary consent, HIPAA Authorization for Research EHRs, and Consent for Return of Genomic Results. The protocol was reviewed by the Institutional Review Board (IRB) of the All of Us Research Program. The All of Us IRB follows the regulations and guidance of the NIH Office for Human Research Protections for all studies, ensuring that the rights and welfare of research participants are overseen and protected uniformly.

Funding

The All of Us program is supported by federal appropriations from Congress. Its primary sources of funding to date have been the Cures Act and a base appropriation allocated to the NIH Office of the Director, which has helped buffer variations in Cures Act funding. The program has historically benefited from strong bipartisan backing and consistent annual funding.

### **Australian Genetics of Depression Study (AGDS)** | Byrne EM, et al. 2020 (PMID 32461290)

Cohort Description

The Australian Genetics of Depression (AGDS) study was established to recruit a large cohort of individuals who have been diagnosed with depression at some point in their lifetime. The purpose of establishing this cohort is to investigate genetic and environmental risk factors for depression and response to commonly prescribed antidepressants. Over 22,000 participants were recruited for AGDS between 2016 and 2018 via Australian government prescription records or through a media campaign. Participants completed online questionnaires within several modules, which consisted of a core module (primarily assessing depression, essential information on self-report mental health diagnoses, medication response and side effects) and 10 ten additional 'satellite' modules that assessed a range of complex traits of relevance to mental health using a variety of scales and questionnaires. Substance use was one of these modules, which was completed by ~73% of AGDS participants (N = ~15,200). Full details about the AGDS cohort and data are published elsewhere (PMID 32461290).

Phenotype Measure

As part of the larger substance use module questionnaire, a range of different measures assessing past, present, and lifetime cannabis were collected, along with diagnostic data for tobacco, alcohol, and other illicit substances. Frequency of cannabis use in the lifetime was assessed by the question: "How many times in your life have you used cannabis (marijuana)?" [responses: Fewer than 5 times; Between 5 and 9 times; Between 10 and 19 times; 20 times or more]. Maximum frequency of cannabis use was assessed with the item: "During the period that you used cannabis the most, how often did you use it? (Non-medical use only: do not include items that were taken in quantities and manner prescribed by a medical professional)" [responses: Once or Twice; Monthly; Weekly; Daily or almost daily]. Additionally, frequency of cannabis use in the past three months was assessed with the item: "In the past three months, how often have you used cannabis (marijuana)? (Non-medical use only: do not include items that were taken in quantities and manner prescribed by a medical professional)" [responses: Never; Once or Twice; Monthly; Weekly; Daily or almost daily]. The average age of participants responding to these questions was 40 years old (SD = 13).

Genotyping, QC, Imputation

Approximately 17,000 of all AGDS participants provided a saliva sample for genotyping (Isohelix GeneFix GFX-02 2 mL saliva collector). Genotyping was conducted using the Illumina Global Screening Array V.2.0 (GSA). Genotype calling was performed with GenomeStudio, and imputation was performed on the Michigan Imputation Server using the HRC reference panel (imputation was run on all markers which pass QC in all batches). The following variant-level quality control was implemented: markers were excluded if there was unknown or ambiguous map position / strand alignment, missingness was greater than 5%, violation of Hardy-Weinberg Equilibrium ($p < 1x10^{-6}$), minor allele frequency less than 1%, and GenTrain score less than 0.6 (if available for that batch). The following individual-level quality control was performed: individuals were excluded if missingness was greater than 3%, sex was incorrect / mismatched and could not be resolved (e.g. could not be explained by sample mix-up, or by an identifiable error in our database), and there were sample duplicates that could not be resolved.

Ethics Statement

All protocols and questionnaires for the AGDS cohort were approved by the QIMR Berghofer Medical Research Institute Human Research Ethics Committee.

Funding
The Australian Genetics of Depression Study was primarily funded by grant 1086683 from the NHMRC of Australia. This work was further supported by NHMRC grants 1145645, 1078901, and 108788, and by National Institutes of Health grant 1R01MH121545-01.

### **Avon Longitudinal Study of Parents and Children (ALSPAC)** | Timpson NJ | PMID 22507743, 31020050, 22507742

Cohort Description

Pregnant women resident in Avon, UK with expected dates of delivery between 1st April 1991 and 31st December 1992 were invited to take part in the study. 20,248 pregnancies have been identified as being eligible and the initial number of pregnancies enrolled was 14,541. Of the initial pregnancies, there was a total of 14,676 fetuses, resulting in 14,062 live births and 13,988 children who were alive at 1 year of age. When the oldest children were approximately 7 years of age, an attempt was made to bolster the initial sample with eligible cases who had failed to join the study originally. As a result, when considering variables collected from the age of seven onwards (and potentially abstracted from obstetric notes) there are data available for more than the 14,541 pregnancies mentioned above: The number of new pregnancies not in the initial sample (known as Phase I enrolment) that are currently represented in the released data and reflecting enrolment status at the age of 24 is 906, resulting in an additional 913 children being enrolled (456, 262 and 195 recruited during Phases II, III and IV respectively). The phases of enrolment are described in more detail in the cohort profile paper and its update update (Boyd et al., 2013; Fraser et al., 2013; Northstone et al., 2019). The total sample size for analyses using any data collected after the age of seven is therefore 15,447 pregnancies, resulting in 15,658 fetuses. Of these 14,901 children were alive at 1 year of age. Study data were collected and managed using REDCap electronic data capture tools hosted at the University of Bristol (Harris et al., 2009). REDCap (Research Electronic Data Capture) is a secure, web-based software platform designed to support data capture for research studies

Phenotype Measure

A binary variable indicating use of cannabis at frequency of once or twice/less than monthly/monthly (0) or weekly/almost daily/daily (1) between ages 13-24 years, using ALSPAC assessments TF1, TF2, TF3, CCR, CCS, TF4, YPB, CCU, CCT and F24. If participants reported different cannabis use frequency at different assessments, the highest reported frequency was used. Due to variation in question at different time points, frequency may refer to past 6 months, 12 months, or time period unspecified (plausibly, current use). If participants had reported the same frequency at multiple assessments, the analysis was adjusted for age at the earliest of these assessments. Please note that the study website contains details of all the data that is available through a fully searchable data dictionary and variable search tool (http://www.bristol.ac.uk/alspac/researchers/our-data/)

Genotyping, QC, Imputation

ALSPAC children were genotyped using the Illumina HumanHap550 quad chip genotyping platforms by 23andme subcontracting the Wellcome Trust Sanger Institute, Cambridge, UK and the Laboratory Corporation of America, Burlington, NC, US. The resulting raw genome-wide data were subjected to standard quality control methods. Individuals were excluded on the basis of gender mismatches; minimal or excessive heterozygosity; disproportionate levels of individual missingness (>3%) and insufficient sample replication (IBD < 0.8). Population stratification was assessed by multidimensional scaling analysis and compared with Hapmap II (release 22) European descent (CEU), Han Chinese, Japanese and Yoruba reference populations; all individuals with non-European ancestry were removed. SNPs with a minor allele frequency of < 1%, a call rate of < 95% or evidence for violations of Hardy-Weinberg equilibrium (P < 5E-7) were removed. Cryptic relatedness was measured as proportion of identity by descent (IBD > 0.1). Related subjects that passed all other quality control thresholds were retained during subsequent phasing and imputation. 9,115 subjects and 500,527 SNPs passed these quality control filters.

The HRC panel was phased using ShapeIt v2, and the imputation was performed using the Michigan imputation server.

Ethics Statement

Ethical approval for the study was obtained from the ALSPAC Ethics and Law Committee and the Local Research Ethics Committees. Informed consent for the use of all data collected was obtained from participants following the recommendations of the ALSPAC Ethics and Law Committee at the time. Participants can contact the study team at any time to retrospectively withdraw consent for their data to be used. Study participation is voluntary and during all data collection sweeps, information was provided on the intended use of data. The completion of a questionnaire, either on paper or online, was considered to be written consent from participants to use their data for research purposes. Biological samples are collected in accordance with the Human Tissue Act (2004). Specific Research Ethics Committee approval is sought for the consenting process at each collection sweep. Written consent, including permission for future use, is obtained from adult participants or from the parents of children as appropriate. Ethical approval for future use is covered by ALSPAC's Research Tissue Bank approval. All historical consents to hold biological samples have been reviewed as part of the Tissue Bank approval process. Participants can contact the study team at any time to retrospectively withdraw consent for use of their samples.

Data Availability

The informed consent obtained from ALSPAC (Avon Longitudinal Study of Parents and Children) participants does not allow the data to be made available through any third party maintained public repository. Supporting data are available from ALSPAC on request under the approved proposal number, B3408. Full instructions for applying for data access can be found here: http://www.bristol.ac.uk/alspac/researchers/access/. The ALSPAC study website contains details of all available data (http://www.bristol.ac.uk/alspac/researchers/our-data/).

Funding

The UK Medical Research Council and Wellcome (Grant ref: 217065/Z/19/Z) and the University of Bristol provide core support for ALSPAC. This publication is the work of the authors and MRM and HMS will serve as guarantors for the contents of this paper. A comprehensive list of grants funding is available on the ALSPAC website (http://www.bristol.ac.uk/alspac/external/documents/grant-acknowledgements.pdf). This research was specifically funded by the following grants from the MRC, Wellcome Trust, and NIH to Matt Hickman, Stan Zammit, George Davey Smith, Glyn Lewis and Ken Kendler (MR/M006727/1, G0800612/86812, 76467/Z/05/Z, 092731/Z/10/Z, 5R01AA018333-05, PD301198- SC101645). GWAS data was generated by Sample Logistics and Genotyping Facilities at Wellcome Sanger Institute and LabCorp (Laboratory Corporation of America) using support from 23andMe.

Acknowledgements

We are extremely grateful to all the families who took part in this study, the midwives for their help in recruiting them, and the whole ALSPAC team, which includes data collection staff, data and administrations staff, technical managers and the technical staff with the Bristol Bioresource Laboratory, based within the University of Bristol.

### **Brisbane Longitudinal Twin Study (BLTS)** | Gillespie NA, Martin NG, Hickie IB | PMID 23245960

Cohort Description

The Brisbane Longitudinal Twin Study (BLTS) is a prospective cohort study that began in 1992 when twins were recruited from primary and secondary schools in the greater Brisbane area via media appeals and by word of mouth. The BLTS cohort consists of both adolescent and young adult twins (3,408 individuals) and their non-twin siblings (1,572), constituting 1,703 families. This includes both monozygotic (MZ) and dizygotic (DZ) twin pairs, including opposite-sex DZ twin pairs, along with singleton siblings of twins, and the twins' parents. The proportions of twins by sex and zygosity in the sample closely mirror population expectations, further strengthening confidence in its representativeness. The BLTS has been recruiting approximately 100 new twins per year and is now a longitudinal collection of psychiatric phenotypes, environmental and psychological risk factors, as well as neurobiological correlates and endophenotypes for psychiatric disorders. The participants are predominantly of European ancestry, reflecting the population structure of Australia at the time of initial recruitment.

Phenotype Measure

As part of the NIH/NIDA "Pathways to Cannabis Use, Abuse and Dependence" project, the study has collected DSM-IV and DSM-V item level data on cannabis abuse and dependence and diagnostic data for nicotine and alcohol, as well as pilot epidemiological data for ecstasy and methamphetamine use. Using the same protocol, the study has also obtained DSM-IV item-level data on mood, anxiety, and fatigue with funding from the NHMRC. The study assessed lifetime use of alcohol, nicotine, cannabis, as well as cocaine, amphetamine-type stimulants, inhalants, sedatives or sleeping pills, hallucinogens, opioids, ecstasy, ketamine, GHB, party drugs, and over-the-counter and prescription analgesics and stimulants for non-medical purposes. In addition to age of initiation, lifetime and past 3-month use for each substance, subjects were also asked if they had ever used or consumed any of the substances while drinking alcohol. For cannabis specifically, the study collected detailed information on lifetime use, frequency of use, age of initiation, age of most frequent use, and quantity consumed during periods of most frequent use. The mental health sections were based on a modified Composite International Diagnostic Interview using item-based criteria to determine caseness for various DSM-IV disorders.

Genotyping, QC, Imputation

All twins and siblings with survey and CATI data have been or will be genotyped using the Illumina 610k SNP array. As of May 2012, approximately 2,639 (74% of the sample) had been genotyped. Extensive quality control has been performed using PLINK. This has included tests of Hardy Weinberg Equilibrium, analysis of missing genotype rates, inbreeding, identity by state, identity by descent statistics for individuals and pairs of individuals, non-Mendelian transmission in family data (when available), sex checks based on X chromosome short nucleotide polymorphisms (SNPs), and tests of non-random genotyping failure. The data were then imputed to contain 2,428,106 SNPs. Imputation boosts the power of many chips toward levels obtained from hypothetical 'complete' arrays containing all HapMap SNPs. Moreover, imputation, which is easily implemented in the software program PLINK, combines information across multiple reference panels, which means that genome-wide association study (GWAS) data obtained from different arrays can be merged for future meta-analyses. A subset of the BLTS subjects have participated in the Brisbane Systems Genetics Study funded by the Australian NHMRC. This included assessment of genome-wide expression on 870 individuals using the Illumina HumanHT-12 v3.0 413 Beadchip, and methylation status on approximately 630 of the same subjects at ~485,000 CpG sites across the genome using the Infinium HM450 and HM27 BeadChips.

Ethics Statement

Protocols for the online survey and CATI were extensively reviewed prior to approval by the QIMR Human Research and Ethics Committee (HREC) and the Virginia Commonwealth University (VCU) Institutional Review Board (IRB). For all studies, written, informed consent was obtained from a parent or guardian and ethics approval was obtained from the Human Research Ethics Committee at the QIMR.

Funding

The "Pathways to Cannabis Use, Abuse and Dependence" project was funded by the US National Institute on Drug Abuse (NIDA) K99R00 award (R00DA023549). Data collection was also supported by the Australian National Health and Medical Research Council (NHMRC) (No. 464914).

### Christchurch Health and Development Study (CHDS) | PIs: David M. Fergusson, L. John Horwood | PMID: 11437801

Cohort Description

The Christchurch Health and Development Study is a longitudinal study of a birth cohort from New Zealand. The cohort was based on an unselected sample of 1,265 consecutive births (635 males; 630 females) occurring in the Christchurch urban region in mid-1977. The cohort has been studied at birth, 4 months of age, 1 year of age, annual intervals to the age of 16 years, and again at ages 18, 21, 25, and 30 years. Sample retention rates were high throughout the study and at age 30 the study was still able to assess over 80% of the surviving cohort. Key areas of data collection included prenatal and perinatal history, family social background, parental characteristics, family change and stability, exposure to child abuse and family dysfunction, child health and health care utilization, educational achievement, behavioral adjustment at school, peer affiliations and relationships, mental health and psychosocial adjustment in adolescence and young adulthood, and participation in tertiary education and the workforce. Assessments also covered nicotine, alcohol, and cannabis use and addiction, as well as measures of family and environmental risk.

Cannabis Analysis Sample: Specific details on the cannabis analysis subsample are not delineated separately for CHDS in the available documentation, but the study contributed to multi-site GEDI analyses on cannabis use. After quality controls across GEDI sites, 2,962 individuals provided over 15,000 total observations, with 747 CHDS participants yielding good quality genotype data. These participants were included in analyses focusing on any cannabis use in the past 3 months, among other substances, as part of gene-environment and gene-development interactions.

Phenotype Measure

The CHDS collected extensive information on individual, family, and community risk for psychopathology, including detailed assessments of drug use, abuse, and dependence (substance use disorders: SUD) and comorbid psychiatric disorders diagnosed using the Diagnostic and Statistical Manual (DSM). For cannabis use specifically, the study assessed nicotine, alcohol, and cannabis use and addiction through repeated measures across adolescence and young adulthood. Phenotypes included measures such as any cannabis use in the past 3 months, with environmental exposures (e.g., stressful life events) centered to the study mean. Data harmonization across GEDI sites involved transforming and recoding variables for comparability, such as using multiple items to estimate substance involvement.

Genotyping, QC, Imputation

Beginning in 2004 (at age 28), participants were asked for consent to provide saliva samples for DNA, with 918 (90% of the surviving cohort) consenting. In 2008-2009, consent for the GEDI multi-site GWAS was obtained from 813 participants, of whom 86% provided peripheral blood samples, 8% provided saliva, and 6% provided buccal swabs (the latter not yielding sufficient quality for genotyping). DNA for the CHDS sample was prepared in New Zealand and sent to the Genotyping Shared Resource at the Mayo Clinic Cancer Center for genotyping using Illumina Human660W-Quad v1 DNA Analysis BeadChips. Quality control was carried out in the Department of Genetics at the University of North Carolina, Chapel Hill. SNPs with missing rate >0.01, minor allele frequency (MAF) <0.05, or extreme deviation (p<10^{-6}) from Hardy-Weinberg equilibrium (HWE) were removed. Subjects with missing rate >0.01 or unusual genome-wide homozygosity (|normalized homozygosity rate| >5) were excluded. Pairwise identical-by-descent (IBD) estimation identified unexpected duplicates and relative pairs. Imputation was performed using MACH, with HapMap3 CEU as the reference (given the predominantly White sample, though some were Maori or mixed). After imputation, over 1,193k total SNP values were available for analysis. Principal components analysis (PCA) extracted five components to control for ancestral and cryptic population stratification, using 77,155 to 79,517 independent SNPs. After quality control checks, good quality data were obtained on 747 participants.

Ethics Statement

Participants in all GEDI studies gave consent for their DNA to be genotyped. CHDS subjects gave consent for genotyping only, separate from consent for the rest of the study. The New Zealand government does not permit the data to be deposited in dbGaP. The Institutional Review Boards at all data gathering sites approved this study, and written consent was obtained from all participants.

Funding

This research was supported by the National Institute on Drug Abuse (R01DA024413), the Health Research Council of New Zealand, the National Child Health Research Foundation, the Canterbury Medical Research Foundation, and the New Zealand Lottery Grants Board.

### **Collaborative Study on the Genetics of Alcoholism (COGA)** | D. Dick | PMID: 28073157 & PMC6875768

Cohort Description

The Collaborative Study on the Genetics of Alcoholism (COGA) is a multi-site, family-based study designed to investigate the genetic contributions to alcohol use disorder and related traits. Probands (i.e., index individuals) were identified through alcohol treatment programs at seven U.S. sites. Probands and their families were invited to participate if the family was sufficiently large with two or more members in the COGA catchment areas. Comparison families were recruited from the same communities. The COGA sample includes both European Ancestry (EA) and African Ancestry (AA) participants, making it an ancestrally diverse cohort. The study encompasses multiple generations of families densely affected by alcohol use disorder, providing a valuable resource for examining genetic and environmental influences on alcohol and substance use behaviors.

Phenotype Measure

As part of a comprehensive assessment, COGA collected data on cannabis use patterns among participants. Among subjects who ever used cannabis (N=11,275), measures included lifetime frequency of cannabis use and days per month of use at the period of most frequent use. The frequency data was collected across different racial groups, including White (N=7,882), Black (N=2,884), Asian (N=79), and Other (N=424) participants. For the total sample, the mean lifetime frequency was 1807.16 (SD=3305.51) with a median of 100 occasions, while the mean days per month at most frequent use was 22.49 (SD=10.01) with a median of 30 days. These detailed measures allow for examination of patterns of cannabis use across different racial/ethnic groups within the cohort.

Genotyping, QC, Imputation

Several genotyping arrays were used in the COGA study: the Illumina 1M, Illumina OmniExpress 12VI, Illumina 2.5M (Illumina, San Diego, CA), and Smokescreen (BioRealm LLC, Walnut, CA). Quality control and imputation procedures are described in Lai et al. (2019). For analyses involving parent-offspring trios, imputed genotypes were used in combination with summary statistics from independent GWAS discovery samples to construct genome-wide polygenic risk scores. This approach used ancestry-specific GWAS weights paired with linkage disequilibrium information from ancestry-matched external reference panels.

Ethics Statement

The Institutional Review Boards at all data gathering sites approved this study, and written consent was obtained from all participants. This ensures the ethical conduct of research involving human subjects according to established guidelines and regulations.

Funding

The Collaborative Study on the Genetics of Alcoholism (COGA) is supported by NIH Grant U10AA008401 from the National Institute on Alcohol Abuse and Alcoholism (NIAAA) and the National Institute on Drug Abuse (NIDA). Additional support for specific projects may come from other funding sources, as indicated in the associated publications.

**Estonian Biobank (EstBB)** | Lehto, K; Estonian Biobank Research Team | PMID: 40188112; 38381979.

Cohort Description

The study sample was drawn from the population-based Estonian Biobank (EstBB), which includes over 212,000 volunteers representing approximately 20% of the adult population in Estonia. The EstBB integrates multiple data layers, including genotype and other omics data, electronic health records, and comprehensive information from questionnaires. A detailed description of the EstBB has been published recently (PMID: 40188112).

Phenotype Measure

We leveraged self-reported data from two EstBB questionnaires: Participant baseline questionnaire II (PBQ-II), which was filled out at recruitment as of 2017, and the Mental health online study (MHoS) questionnaire (PMID: 38381979), which was conducted in spring 2021. Both questionnaires included items about lifetime and current illicit drug use. Cases were defined as individuals who 1) responded "Yes" to the PBQ-II item "Have you used narcotic substances other than alcohol and tobacco" and specified any cannabis product in the following free text box (e.g., marijuana, weed, hash, grass) or 2) responded "Yes" to the MHoS questionnaire item "In the past 3 months, did you use cannabis?". Controls were defined as individuals who 1) responded "No" to the PBQ-II item "Have you used narcotic substances other than alcohol and tobacco" and 2) responded "Never" to the MHoS questionnaire item "Have you ever used the following substances regularly?: Narcotics (e.g., cannabis, amphetamine, cocaine, fentanyl, LSD, etc.)" and 3) responded "No" to the MHoS questionnaire item "In the past 3 months, did you use cannabis?".

Genotyping, QC, Imputation

Estonian Biobank participants were genotyped at the Genotyping Core Facility of the Institute of Genomics, University of Tartu, using four revisions of the Illumina GSA array (GSAMD-24v1, GSAMD-24v2, ESTchip-1_GSAv2, ESTchip-2_GSAv3) across multiple genotyping batches, each containing at least 1,000 samples. All quality control (QC) procedures were performed separately for each batch. At the sample level, individuals with a call rate <95% and/or a sex mismatch between genotype and phenotype data were excluded. At the variant level, genotypes with call rates <95% were removed. Variants with poor Illumina cluster separation (<0.4) and/or a GenTrain score <0.6 were also excluded. To control for potential genotyping batch effects, variants showing inconsistent allele frequencies across batches were removed. Specifically, variant allele frequencies were calculated for each genotyping batch containing more than 10,000 samples (9 batches in total). The mean allele frequency across these batches was then computed, and variants with frequencies deviating by more than 5% from the mean in any batch were excluded from the combined dataset. Only single-nucleotide variants (SNVs) were retained for imputation. Among these, AT, GC, multiallelic, and rare variants (minor allele frequency <1%) were removed. After QC, all genotyping batches were merged into a single cohort dataset. Approximately 310K SNVs passed all QC filters and were used for imputation. The dataset was phased reference-free using Eagle v2.4.1. Subsequent imputation of the pre-phased data was performed with Beagle v5 (beagle.22Jul22.46e.jar), using the local copy of the Haplotype Reference Consortium (HRC) reference panel (https://www.sanger.ac.uk/collaboration/haplotype-reference-consortium/), which included 27,165 reference samples and only polymorphic sites with at least 5 minor alleles. For ancestry analysis, EstBB samples were merged with the 1000 Genomes Project reference panel (n = 2,495). Principal component analysis was performed using the *bigsnpr* package (PMID: 35604078), and individuals that were not of European ancestry were removed. The GWAS analysis was conducted using Regenie v3.2. that accounts for population structure through a genetic relatedness matrix. Association analysis with standard binary trait settings was carried out for all variants with an INFO score >0.4 with sex, age at phenotypic assessment, birth cohort, and 20 PCs as covariates.

Ethics Statement

The activities of the EstBB are regulated by the Human Genes Research Act, which was adopted in 2000 specifically for the operations of the EstBB. Individual level data analysis in the EstBB was carried out under ethical approvals [1.1-12/624 and 1.1-12/2860] from the Estonian Committee on Bioethics and Human Research (Estonian Ministry of Social Affairs), using data according to release application [6-7/GI/11576] from the Estonian Biobank.

Funding

This work in EstBB was primarily supported with grant from Estonian Research Council (grant no PSG615). KL and KK were supported by the Estonian Centre of Excellence for Well-Being Sciences, funded by grant TK218 from the Estonian Ministry of Education and Research. The research was conducted using the Estonian Center of Genomics/Roadmap II funded by the Estonian Research Council (project number TT17).

Acknowledgements

We thank all participants and staff of the Estonian Biobank for their contribution to this research. We would also like to thank the Estonian Biobank research team, including Andres Metspalu, Lili Milani, Tõnu Esko, Reedik Mägi, Mari Nelis and Georgi Hudjashov who were responsible for data collection, genotyping, QC and imputation of the EstBB data. Data analysis was carried out in part in the High-Performance Computing Center of University of Tartu.

### **Healthy Life in an Urban Setting (HELIUS)** | Verweij KJH; Zwinderman K | PMID 23621920, 29247091

Cohort Description

Healthy Life in an Urban Setting (HELIUS) is a prospective population-based cohort study executed in Amsterdam, characterized by ethnic diversity. HELIUS includes six large groups of inhabitants of Amsterdam, namely, Dutch, African Surinamese, South-Asian Surinamese, Turkish, Moroccan, or Ghanaian background, and one small group with a Javanese Surinamese background. The HELIUS cohort consists of approximately 25,000 participants aged 18–70 years. For most participants, data on social, environmental, and biological determinants were collected, and follow-up data were obtained. Detailed information on the cohort participants and gathered data has previously been published (PMID 40483003; 23621920; 29247091).

Phenotype Measure

At the baseline data collection, self-report measures of lifetime cannabis use have been collected, along with diagnostic data for nicotine, alcohol, and other illicit substances. The average age at inclusion is 43.8 years (SD=3.65, range=18-70 years). Lifetime cannabis use was assessed by asking: "In your life, have you ever used cannabis (marijuana, pot, grass or hash)?"

Genotyping, QC, Imputation

A cross-selection of 10,285 HELIUS participants was made for genotyping. Whole blood for DNA isolation was collected in EDTA tubes and stored at −80°C in the AMC Biobank. DNA was isolated using the Gentra Puregene Isolation Kit (Qiagen), and quality control procedures were performed to determine the DNA yield and purity. Genotyping was performed at the Erasmus MC Human Genomic Facility, using the Illumina Global Screening Array 24v1-0 designed for the multiethnic genome-wide content purpose was used. An in-house protocol of the Human Genomic Facility, with Illumina's GenomeStudio software, was used to perform the initial genotyping of the array. Subsequently, a second quality control (QC) was performed for removing the individuals with discordant gender information and when more than 5% called data on markers per individual were missing.

Before imputation a general QC for the autosomal markers was executed removing variations with more than 5% calls missing, minor allele frequencies (MAF) of <1%, violation of the Hardy–Weinberg equilibrium (HWE) ($p \leq 10{-}5$), and heterozygosity deviations from a mean larger than ± 3 SD.

Imputation was conducted with the Sanger imputation server using the 1000 Genome phase 3 cohort. All markers are reported with respect to the reference allele and coordinates of GRCh37.

Ethics Statement

The HELIUS study has been approved by the Ethical Review Board of the Amsterdam UMC, location AMC. All participants approved by giving written informed consent.

Funding

Amsterdam UMC, location AMC, and the Public Health Service of Amsterdam (GGD Amsterdam) provided core financial support for HELIUS. The HELIUS study is also funded by research grants of the Dutch Heart Foundation (Hartstichting; grant no. 2010T084), the Netherlands Organization for Health Research and Development (ZonMw; grant no. 200500003), the European Integration Fund (EIF; grant no. 2013EIF013) and the European Union (Seventh Framework Programme, FP-7; grant no. 278901).

### **Finnish Twin Cohort (FinnTwin)** | Ollikainen M | PMID 31640839, 31796134, 29532581

Cohort Description

FT12:

FinnTwin12 was designed as a two-stage study. In the first stage, we conducted multi wave questionnaire research enrolling all eligible twins born in Finland during 1983-1987 along with their biological parents. In stage 2, we intensively studied a subset of these twins with in-school assessments at age 12 and semi structured poly-diagnostic interviews at age 14. At baseline, parents of intensively studied twins were administered the adult version of the interview. Laboratory studies with repeat interviews, neuropsychological tests, and collection of DNA were made of intensively studied twins during follow-up in early adulthood. The basic aim of the FT12 study design was to obtain information on individual, familial and school/neighborhood risks for substance use/abuse prior to the onset of regular tobacco and alcohol use and then track trajectories of use and abuse and their consequences into adulthood. But the longitudinal assessments were not narrowly limited to this basic aim, and with multi wave, multi rater assessments from ages 11 to 12, the study has created a richly informative data set for analyses of gene-environment interactions of both candidate genes and genome wide measures with measured risk-relevant environments. To date, five waves of data collection have been completed.

FT16:

FinnTwin16 was initiated in 1991 to identify the genetic and environmental precursors of alcoholism, but later the scope of the project expanded to studying the determinants of various health-related behaviors and diseases in different stages of life. The main areas addressed are alcohol use and its consequences, smoking, physical activity, overall physical health, eating behaviors and eating disorders, weight development, obesity, life satisfaction and personality. To date, six waves of data collection have been completed.

NAG:

The study sample was drawn from the population-based Finnish Twin Cohort Study, which consists altogether of 35 834 adult twins born in 1938–1957. Twin pairs concordant for ever-smoking were identified and recruited along with their family members (mainly siblings) for the Nicotine Addiction Genetics (NAG) Finland study. Priority was given to heavier smokers. The data collection took place in 2001–2005. Participants were assessed by DNA sample collection and a structured diagnostic psychiatric interview resulting in detailed phenotypic information on multiple smoking behavior traits.

Phenotype Measure

Survey question for lifetime frequency of cannabis use: “How many times have you used cannabis?”; for frequency of cannabis in the past year: “How many times have you used cannabis in the past year?”.

Genotyping, QC, Imputation

Chip genotyping were done using Illumina Human610-Quad v1.0 B, Human670-QuadCustom v1.0 A, Illumina HumanCoreExome- (12 v1.0 A, 12 v1.1 A, 24 v1.0 A, 24 v1.1 A, 24 v1.2 A) and Affymetrix FinnGen Axiom arrays. The algorithm for genotype calling were Illumina’s GenCall for all HumanCoreExome chip genotypes, Illuminus for 610k & 670k chip genotypes and AxiomGT1 for Affymetrix chip genotypes. On Illumina arrays where genotypes were called to Illumina’s TOP strand, strand were flipped to forward strand using strand files generated by Will Rayner (https://www.well.ox.ac.uk/~wrayner/strand/). The genome build of all genotypes were set to GRCh37/hg19. In case where genotypes were called to NCBI36/hg18 or GRCh38/hg38, genome positions were lifted to GRCh37/hg19 using University of

California Santa Cruz LiftOver program [1] with appropriate chain file. Genotype quality control, pre-phasing and imputation were done in three batches (batch1: 610k+670k, batch2: HumanCoreExome and batch3: Affymetrix chip genotypes). We removed variants with call rate below 97,5% (batch1 and batch3) or 95% (batch2), samples with call rate below 98% (batch1) or 95% (batch2 and batch3), variants with minor allele frequency below 1% with Hardy-Weinberg Equilibrium p-value lower than 1e-06. Also samples from all batches with heterozygosity test method-of-moments F coefficient estimate value below -0.03 or higher than 0.05 (batch1 and batch2) or ±4SD from the mean (batch3) were removed along with the samples which failed sex check or were among the multi-dimensional scaling principal component analysis outliers. Total amount of genotyped autosomal variants after quality control were 475526 (batch1), 239894 (batch2) and 388673 (batch3) with following number of samples remaining for imputation: 2617 (batch1), 5328 (batch2) and 8218 (batch3).

We then performed pre-phasing using Eagle v2.3 [2] and imputation with Minimac3 v2.0.1 using University of Michigan Imputation Server [3]. Genotypes of all batches were imputed to Haplotype Reference Consortium release 1.1 reference panel [4]. After imputation three imputation batches were merged and the final study subset were extracted.

1: Hinrichs et al. The UCSC Genome Browser Database: update 2006. Nucleic Acid Research 2006
2:Loh et al. Reference-based phasing using the Haplotype Reference Consortium panel. Nature Genetics 2016.
3: Das et al. Next-generation genotype imputation service and methods. Nature Genetics 2016
4: the Haplotype Reference Consortium. A reference panel of 64,976 haplotypes for genotype imputation. Nature Genetics 2016.

Ethics Statement

The study protocol was approved by the Institutional Ethics Board of the Hospital District of Helsinki and Uusimaa, Finland. All applicable written and informed consent was obtained in relation to the data generated or used for analysis.

Funding

Phenotyping and genotyping of the Finnish twin cohorts was supported by the Academy of Finland Centers of Excellence in Complex Disease Genetics (grants 213506, 129680 and 312073 ), NIDA (grant number R01 DA012854, PI: Pamela A. F. Madden), Sigrid Juselius Foundation (to Jaakko Kaprio), Global Research Award for Nicotine Dependence, Pfizer Inc. (to Jaakko Kaprio), and the Welcome Trust Sanger Institute, UK.

### **Great Smoky Mountains Study (GSMS)** | PIs: William E. Copeland, formerly E. J. Costello, | PMID: 23461817, PMCID: PMC3609892

Cohort Description

The Great Smoky Mountains Study is a longitudinal, population-based study of the development of psychiatric disorders and substance use in rural youth. Three cohorts of children, aged 9, 11, and 13 at intake in 1993, were selected from a rural population of approximately 20,000 children using a household equal probability design. A two-phase procedure was used for White and African-American youth to increase statistical power by oversampling children at risk for psychiatric and substance use disorders. Half of the sample are female, and 6% are African American. American Indian youth were oversampled (100%) as an understudied group known to be at high risk. The total GSMS sample is 1,420 participants. Data collection is complete for ages 9-26, with 9,858 interviews completed by age 26, and age 30 interviews in progress at the time of documentation.

Phenotype Measure

The GSMS used the Child and Adolescent Psychiatric Assessment (CAPA) and its adult version to assess psychiatric symptoms and substance use. For cannabis use specifically, the study collected data on lifetime frequency and days per month during periods of most frequent use.

Genotyping, QC, Imputation

Blood samples were collected using finger-stick methodology at each assessment. Of the GSMS participants, 784 had adequate genotype data and consent to deposit data in dbGaP. All samples were genotyped using Illumina Human660W-Quad v1 DNA Analysis BeadChips. Quality control was carried out in the Department of Genetics at the University of North Carolina, Chapel Hill. Across all four GEDI sites (including VTSABD), 2,962 individuals provided over 15,000 total observations after quality controls.

Ethics Statement

The Institutional Review Boards at all data gathering sites approved this study, and written consent was obtained from all participants. In Year 1 of the GEDI study, additional consent was obtained for depositing biological samples and genetic data in controlled-access biorepositories (e.g., dbGaP).

Funding

This research was supported by the National Institute on Drug Abuse (U01DA024413, R01DA11301), the National Institute of Mental Health (R01MH063970, R01MH063671, R01MH048085, K01MH093731 and K23MH080230), NARSAD, and the William T. Grant Foundation.

### **Lifelines population cohort (LL)** | Debbie van Baarle | PMID 25502107, 34897450

Cohort Description

The Lifelines Cohort Study is a large population-based cohort study and biobank that was established as a resource for research on complex interactions between environmental, phenotypic and genomic factors in the development of chronic diseases and healthy ageing. Between 2006 and 2013, inhabitants of the northern part of the Netherlands and their families were invited to participate, thereby contributing to a three-generation design. Participants visited one of the Lifelines research sites for a physical examination, including lung function, ECG, and cognitive tests, and completed extensive questionnaires. Follow-up visits are scheduled every 5 years, and in between participants receive follow-up questionnaires. Linkage is being established with medical registries and environmental data. Lifelines contains information on biochemistry, medical history, psychosocial characteristics, lifestyle and more.

Phenotype Measures

Cannabis usage was determined based on questionnaire data. During the second assessment, participants were asked the following questions:
Have you ever used any of the drugs listed below? (weed, marijuana, hashish)
How often did you use drugs in the past 12 months? (weed, marijuana, hashish)
How often did you use drugs in your entire life? (weed, marijuana, hashish)
Note that the frequency questions were multiple-choice, with the possible answers: 0, 1, 2, 4, 6, 11-19, 20-39, and 40+.

Genotyping, QC, Imputation:

Lifelines was genotyped in two batches, which were imputed separately. The first batch used the Illumina Human Cyto SNP12 v2; the second the Illumina Global Screening Array (GSA). Both batches were imputed to HRC on the Sanger Imputation server. Analysis was performed using SAIGEgds.

Ethics Statement:

The Lifelines study was approved by the ethics committee of the University Medical Center Groningen, document number METC UMCG METc 2007/152. Informed consent was obtained from all individuals included in the study.

Funding

The Lifelines Biobank initiative has been made possible by funding from the Dutch Ministry of Health, Welfare and Sport, the Dutch Ministry of Economic Affairs, the University Medical Center Groningen (UMCG the Netherlands), University of Groningen and the Northern Provinces of the Netherlands. The generation and management of GWAS genotype data for the Lifelines Cohort Study is supported by the UMCG Genetics Lifelines Initiative (UGLI). UGLI is partly supported by a Spinoza Grant from NWO, awarded to Cisca Wijmenga.

### Minnesota Center for Twin and Family Research (MCTFR) Study | Principal Investigators: Matt McGue, William G. Iacono | PMID: 22877857, 23897083, 23314086

Cohort Description

The Minnesota Center for Twin and Family Research (MCTFR) is a collaborative research initiative within the Gene-Environment-Development Initiative (GEDI), comprising three longitudinal studies: 1) the Minnesota Twin Family Study (MTFS), 2) the Enrichment Sample (ES), and 3) the Sibling Interaction and Behavior Study (SIBS). The MTFS includes two intake cohorts of same-sex twins, one assessed initially at age 11 (Cohort 1) and another at age 17 (Cohort 2), identified through Minnesota birth records. The ES consists of same-sex twins screened for elevated symptoms of attention-deficit/hyperactivity disorder (ADHD) and conduct disorder, first assessed at age 11. The SIBS is an adoption study involving adopted individuals, their unrelated adoptive siblings, and parents. The current GWAS sample for cannabis use frequency comprises 7,188 European-ancestry individuals (3,344 males, 4,087 females) clustered in approximately 2,300 families, consisting of twin pairs, siblings, and their rearing parents.

Phenotype Measures

Cannabis use frequency was assessed via semi-structured interviews based on the Composite International Diagnostic Interview - Substance Abuse Module (CIDI-SAM). Individuals reporting lifetime marijuana use and at least 5 uses since the last assessment received follow-up questions.

Last 12 months frequency (n = 1,851; 1,073 males, 778 females; mean age = 32.0, SD = 11.1, range 16–59; data collected 1990–2015). Question: “How frequently have you used marijuana in the last 12 months?”
Maximum lifetime frequency (n = 2,576; 1,460 males, 1,116 females; mean age = 20.5, SD = 4.6, range 11–48; data collected 1990–2019). Question: “How frequently did you use marijuana during your period of heaviest use?”

Both measures used a 1–10 forced-choice scale (1 = 3+ times/day; 10 = less than once a year), reverse-coded and collapsed to an ordinal 0–7 scale: 0 = not in the last 12 months; 1 = < once/month; 2 = once monthly; 3 = 2–3 times/month; 4 = 1–2 times/week; 5 = 3–4 times/week; 6 = nearly daily; 7 = daily/multiple times daily. Lifetime ever-use prevalence (N=7,431 genotyped European-ancestry individuals): 4,028 ever users, 3,403 never users (data collected 1991–2013; age range 19–65; mean age = 33.27, SD = 11.9).

Genotyping, QC, Imputation

Genotyping was performed using the Illumina Human660W-Quad Array (561,490 SNPs). Quality control excluded markers with call rate <99%, MAF <1%, HWE $p < 10^{-7}$, or other standard criteria, retaining 515,384 autosomal SNPs. Imputation for candidate SNPs used HapMap2 ($r^2 \geq 0.917$). GWAS for last-12-months frequency (N=7,188 pre-QC) was conducted using RFGLS (linear model) in a variance-covariance matrix framework accounting for family structure, adjusting for generation, generation × age, generation × birth year, and 10 ancestry PCs.

Ethics Statement

National Institute on Drug Abuse (NIDA) grant R01DA024413 (Gene-Environment-Development Initiative, GEDI), with additional support from the National Institutes of Health.

Funding

The MCTFR was supported by the National Institute on Drug Abuse (NIDA) under grant R01DA024413 as part of the Gene-Environment-Development Initiative (GEDI), with additional funding from the National Institutes of Health.

### Netherlands Mental Health Survey and Incidence Study – 2 (NEMESIS-2) | ten Have M | PMID 20641046, 21197531

Cohort Description

NEMESIS-2 is a Dutch population-based cohort with adults aged 18-64 years, developed to study the occurrence of mental disorders and wellbeing with face-to-face interviews. It uses a multistage, stratified random sampling of households, with one respondent randomly selected from each household. In the first wave (T0), performed from November 2007 to July 2009, 6,646 individuals were interviewed (response rate 65.1%). This sample was nationally representative, although younger subjects were somewhat underrepresented (De Graaf et al., 2010). All respondents were approached for a follow-up interview at three years (T1: n=5,303; response rate 80.4%), six years (T2: n=4,618; response rate 87.8%) and nine years (T3: n=4,007; response rate 87.7%) after baseline. Attrition between T0 and T3 was not significantly associated with 12-month prevalence of all in the study assessed mental disorders at T0 after controlling for sociodemographic characteristics (De Graaf et al., 2018).

Phenotype Measure

Cannabis use was assessed with the section Illegal Substance Use of the CIDI 3.0 at baseline (T0; lifetime). If subjects reported cannabis use, they were rated on frequency of use in the last 12 months and in the period of most frequent use. The item wording for use in the last 12 months was: “How often did you use marijuana in the past 12 months?”, and for period of most use: “Think back to the year when you used marijuana the most. How often did you use then?”. For both, the response options were (6) every day, (5) almost every day, (4) 3-4 days per week, (3) 1-2 days per week, (2) 1-3 days per month, and (1) less than once per month.

Genotyping, QC, Imputation

NEMESIS-2 samples were genotyped on an IPMCN chip (Institute of Psychological Medicine and Clinical Neurology, Cardiff University), which was custom-made for EUGEI (588,628 genotyped variants for 4,043 participants).[6] Quality control (QC) was done using PLINK v1.9 [7] as follows**.** There were 3,861 samples matching with phenotypes. Single nucleotide polymorphisms (SNPs) and samples with call rates below 95% and 98%, respectively, were removed. A strict SNP QC only for subsequent sample QC steps was conducted. This involved a minor allele frequency (MAF) threshold > 10% and a Hardy-Weinberg equilibrium (HWE) *P*-value > $10^{-5}$, followed by linkage disequilibrium (LD) based SNP pruning ($R^2 < 0.5$). This resulted in ~60K SNPs to assess sex errors (n=145), heterozygosity (F< 5xSD the standard deviation (SD), n= 73), and relatedness by pairwise identity by descent (IBD) values > 0.1 (n= 170). Genetic outliers (n=154) were identified by principal component analysis (PCA, see below). In total, 3,104 individuals passed these QC steps. After removing failing samples (n = 757), a regular SNP QC was performed (SNP call rate>95%, HWE p>1e-06, MAF> 0.16%; as the IPMCN chip contains many rare variants, half the SNPs would have been removed if we had applied MAF> 1%; therefore, we loosened MAF threshold to 0.16% =10/(2*sample size of 3,104)). Next, strand ambiguous SNPs and duplicate SNPs were removed, resulting in a total of 298,104 genotyped variants. The QC-ed dataset was chunked by chromosome, and then converted into *.VCF files. The Michigan server was used for imputation with the following settings: reference panel as HRC R1.1 2016; phasing as Eagle v2.3; population as European; model as QC & imputation. The imputation resulted in 47,101,073 single nucleotide polymorphisms (SNPs). All markers are reported with respect to the reference allele and coordinates of GRCh37.

Ethics Statement

The study was approved by a medical ethics committee (the Medical Ethics Review Committee for Institutions on Mental Health Care, METIGG; reference number: CCMO/NL18210.097.07). After receiving information about the study aims, respondents provided written informed consent at each wave.

Funding

Financial support has been received from the Ministry of Health, Welfare and Sport, with supplementary support from the Netherlands Organization for Health Research and Development (ZonMw) and the Genetic Risk and Outcome of Psychosis (GROUP) investigators. The funding sources had no further role in study design; in the collection, analysis and interpretation of data; in the writing of the report; or in the decision to submit the paper for publication.

### **Nicotine Addiction Genetics (NAG), QIMR**| Martin NG, Madden PAF, Heath ACH | PMID 17436240

Cohort Description
The NAG project was an international collaborative study that included three sites (QIMR Berghofer, University of Helsinki and Washington University). Australian participants were enrolled at QIMR Berghofer (formally the Queensland Institute of Medical Research) in Brisbane, Australia. Families were identified through heavy-smoking index cases by use of previously administered interview and questionnaire surveys of the community-based Australian register of twins. Families were identified from two cohorts of the Australian Twin Panel, which included spouses of the older of these two cohorts, for a total of ~12,500 families with information about smoking. The ancestry of the Australian samples is predominantly Anglo-Celtic or northern European (>90%).

Phenotype Measure
As part of a larger study, participants were interviewed using the diagnostic Semi-Structured Assessment for the Genetics of Alcoholism (SSAGA) protocol. The average age at interview was 41.1 years (SD=7.3, range=18-73). Lifetime cannabis use was assessed by asking "How many times in your life have you used marijuana?". Due to extreme skewness of responses, the data was assigned to six bins: 1=Once or Twice; 2=3-5 times; 3=6-12 times, 4=13-100 times, 5=101-500 times, 6=More than 500 times. Participants were also assessed for their most frequent cannabis use by asking "Think about the period of time when you were using marijuana the most. During that period of time, how often were you using marijuana?". Data kurtosis was reduced using log10 transformation.

Genotyping, QC, Imputation
Participant gave informed consent for providing a blood samples for DNA extraction. Samples were genotyped on Illumina platforms, including the Illumina CNV370-Quadv3 platform (Illumina, Inc.) by the Center for Inherited Disease Research (CIDR, Baltimore, Maryland, USA) and by deCODE (Reykjavik, Iceland), the Illumina 317K platform by the University of Helsinki Genome Center (Helsinki, Finland), and the Illumina 610 Quad platform by deCODE. Genotypes were called using Illumina BeadStudio software. Quality-control excluded SNPs with mean GenCall score less than 70%, with call rate less than 95%, with deviation from Hardy-Weinberg significant at $p<10^{-6}$, or Minor Allele Frequency less than 1%. Imputation was conducted using the Michigan imputation server using the 1000 Genome phase 3 reference panel. All markers are reported with respect to the reference allele and coordinates of GRCh37.

Ethics Statement
Informed consent was obtained from participants and the study was approved by the Institutional Review Boards at Washington University School of Medicine, the University of Missouri, and the Human Research Ethics Committee of QIMR Berghofer.

Funding
The NC study was supported by National Institutes of Health grants DA12854, AA07728, AA07580, AA13321, AA13320, and DA019951. Funding was also provided by the Australian National Health and Medical Research Council (241944, 339462, 389927, 389875, 389891, 389892, 389938, 442915, 442981, 496739, 552485, 552498). A portion of the genotyping on which this study was based was carried out at the Center for Inherited Disease Research, Baltimore (CIDR), through an access award to our late colleague Dr. Richard Todd.

### **The Netherlands Twin Register (NTR)** | Boomsma DI | PMID 31666148

Cohort Description

The Netherlands Twin Register (NTR) is a population-based prospective cohort study which includes newborn twins and multiples from the Netherlands. Recruitment started with birth year 1986. NTR data collection has a focus on growth, development, emotional and behavioral problems and health. More information may be found at: https://tweelingenregister.vu.nl/nl.

Phenotype Measure

Phenotype data on cannabis use were collected by self-report surveys, The average age at interview is 34.55 years (SD=12.21, range= 18-65). Cannabis use was assessed by asking: "*How often in your life did you use cannabis*?", "*How often did you use cannabis in the period you used the heaviest?", and "Did you use cannabis in the past year?"*

Genotyping, QC, Imputation

Buccal cells and blood for DNA isolation were collected in multiple sub-projects. Quality control and imputation were performed on a larger NTR dataset (N= 23,601). SNP data were cleaned with the following criteria: Hardy–Weinberg equilibrium *p* value >.00001, minor allele frequency >0.01 & <0.4, call rate >0.95, Mendelian errors <21, and Mendelian error rate < mean + 3 sd. Samples were cleaned with the following criteria: call rate >0.90, Plink heterozygosity −0.10 < F < 0.10, and consistency of X chromosome genotypes with known gender. Imputation was conducted using an Haplotype Reference Consortium (HRC) reference panel. All markers are reported with respect to the reference allele and coordinates of GRCh37.

Ethics Statement

The study was approved by the Central Ethics Committee on Research Involving Human Subjects of the VU University Medical Centre, Amsterdam, an Institutional Review Board certified by the U.S. Office of Human Research Protections (IRB number IRB00002991 under Federal-wide Assurance- FWA00017598; IRB/institute codes, NTR 03-180).

Funding

Funding was obtained from multiple grants from the Netherlands Organization for Scientific Research (NWO) and The Netherlands Organisation for Health Research and Development (ZonMW): Genetic influences on stability and change in psychopathology from childhood to young adulthood (ZonMw 912-10-020); Twin family database for behavior genomics studies (NWO 480-04-004); Genetic and Family Influences on Adolescent. Psychopathology and Wellness (NWO 463-06-001); A Twin-Sibling Study of Adolescent Wellness (451-04-034)**.** Twin research focusing on behavior (NWO 400-05-717); Longitudinal data collection from teachers of Dutch twins and their siblings (481-08-011), Twin-family-study of individual differences in school achievement (NWO-FES, 056-32-010), Genotype/phenotype database for behavior genetic and genetic epidemiological studies (ZonMw Middelgroot 911-09-032); "Why some children thrive" (OCW_Gravity program –NWO-024.001.003), Netherlands Twin Registry Repository: researching the interplay between genome and environment (NWO-Groot 480-15-001/674); BBMRI –NL (184.021.007 and 184.033.111): Biobanking and Biomolecular Resources Research Infrastructure; Spinozapremie (NWO- 56-464-14192) and KNAW Academy Professor Award (PAH/6635) to DIB ; the Neuroscience Campus Amsterdam (NCA) and Amsterdam Public Health (APH); the European Science Council (ERC) Genetics of Mental Illness (ERC Advanced, 230374); NIH: Rutgers University Cell and DNA Repository cooperative agreement (NIMH U24 MH068457-06); Developmental Study of Attention

Problems in Young Twins (NIMH, RO1 MH58799-03); Grand Opportunity grant Developmental trajectories of psychopathology (NIMH 1RC2 MH089995) and the Avera Institute for Human Genetics.

### **Spit for Science (S4S)** | Dick DM, Kendler KS | PMID 24639683, PMID 29247091

Cohort Description

Spit for Science (S4S) is a university-wide longitudinal study at Virginia Commonwealth University (VCU) aimed at understanding how genetic and environmental influences impact alcohol use and related substance use and mental health outcomes across time in college students. The study began in 2011 with cohorts of freshman participants enrolled each year from 2011 to 2014. Nearly 70% of the incoming freshman class (N=2,715) completed online surveys in the first wave, with 80% of the students completing spring follow-ups. The sample closely approximated the university population in terms of gender and racial/ethnic composition, with 60% female and 40% male participants. The racial distribution was 15% Asian, 20% African American, 7% Hispanic, 6% more than one race, and 50% White. Participants were followed annually with surveys each spring throughout their college years. This longitudinal design, combined with genetic data collection, allows for the study of genetic and environmental influences on substance use trajectories across this critical developmental period.

Phenotype Measure

Cannabis use was assessed in Spit for Science through two primary measures: lifetime cannabis use and past 12-month cannabis use. For lifetime cannabis use, participants were asked "How many times have you ever used cannabis?" with a free response format. For recent use, participants were asked "How many times have you used cannabis over the last 12 months?" also with a free response format. These items were part of a larger battery of substance use questions assessing various licit and illicit drugs. The survey also included detailed assessment of alcohol use and problems, including DSM-IV based criteria to assess alcohol abuse and dependence. Various forms of social activity participation, peer deviance, and mental health outcomes were also measured, allowing for comprehensive analysis of risk and protective factors for substance use.

Genotyping, QC, Imputation

DNA samples were collected from saliva samples provided by 98% of eligible participants. Four milliliters of saliva were collected from each participant in Oragene collection tubes and DNA was isolated following manufacturer's instructions. Genotyping was performed at Rutgers University Cell and DNA Repository (RUCDR) using the Affymetrix BioBank array. Pre-imputation quality control removed Off Target Variants identified in SNPolisher, SNPs missing more than 5% of genotypes, samples missing more than 2% of genotypes, and SNPs missing more than 2% of genotypes after sample filtering. Imputation was conducted using the 1000 Genomes phase 3 reference panel. Ancestry was empirically assigned using SmartPCA (Eigenstrat) to match each DNA sample to the best fitting 1000 Genomes reference population using Mahalanobis distance. For construction of polygenic scores, SNPs were clumped by linkage disequilibrium (LD) using the clump procedure in PLINK, based on an $R^2 = 0.25$ and 500kb window. For European American samples, this resulted in 1,308,830 SNPs for creating polygenic risk scores, and 1,307,510 SNPs for African American samples.

Ethics Statement

The Spit for Science study was approved by the Virginia Commonwealth University Institutional Review Board. All participants provided informed consent for participation in both the survey and DNA components of the study. Extensive groundwork was conducted before launching the project to develop support, raise awareness, and minimize misunderstanding that could surround a large-scale university-wide study with a genetic component. This included meeting with multiple stakeholders across the university and developing informational materials about the study and study-specific website. The research team also worked with the university's Wellness Resource Center to develop informational materials about alcohol use on college campuses to mail to parents.

Funding

Spit for Science was funded by R37AA011408 from the National Institute on Alcohol Abuse and Alcoholism (NIAAA) to Kenneth S. Kendler, with support for Danielle M. Dick through K02AA018755. Additional support for the project was obtained through National Institutes of Health P20 AA107828, Virginia Commonwealth University, and UL1RR031990 from the National Center for Research Resources and National Institutes of Health Roadmap for Medical Research. The cannabis genetic research component was supported by the National Institute on Drug Abuse (NIDA), which funded the "Pathways to Cannabis Use, Abuse, and Dependence" project to explore the genetic and environmental risk factors in the pathways to cannabis use disorder.

### **Saguenay Youth Study (SYS)** | Paus T; Pausova Z| PMID 25454417, 27018016

Cohort Description

The Saguenay Youth Study (SYS) is a population-based study of adolescents and their middle-aged parents1. It is aimed at investigating the etiology, early stages and trans-generational trajectories of common cardio-metabolic and brain diseases. The SYS was designed as a two-generational cohort; it includes 1,029 adolescents and their 962 parents. The cohort was recruited via 12- to 18-year-old adolescents attending high schools in the Saguenay Lac-Saint-Jean region of Quebec (Canada). Half of the adolescents were exposed prenatally to maternal cigarette smoking. The cohort is family-based (n=481 families), including only adolescents who have one or more siblings of similar age (i.e., 12 to 18 years) and both biological parents of the French-Canadian origin born in the region. The data collection occurred in two waves. Wave 1 (2003-2012) involved the recruitment and complete assessment of all 1,028 adolescents, as well as a partial ('soft') assessment of 962 parents. Wave 2 (2012-2015) involved the complete assessment of a subset of the parents (n=664). In Wave 2, parents answered a series of questions about their drug use; this questionnaire was based on the European School Survey Project on Alcohol and Other Drugs (http://www.espad.org/). Data from SYS parent cohort were included in the present study. Detailed information on the SYS participants and gathered data has previously been published (PMID 25454417; 27018016).

Phenotype Measure

Cannabis-use was measured based on responses (Yes/No) to the following question: *"Have you ever used marijuana (grass, pot) or hashish (hash, hash oil)?"*. Of the total sample (N=565), 278 individuals reported never using cannabis use, while 287 individuals reported cannabis use. Participants who answered "Yes" were further asked about the frequency of their lifetime cannabis use. Responses were recorded using the following scale: "1=once or twice; 2=3-5 times; 3=6-9 times; 4=10-19 times; 5=20 to 39 times; and 6= 40 times and more".

Genotyping, QC, Imputation

The SYS adolescents and parents were genotyped in two waves. First, 592 adolescents were genotyped with the Illumina Human610-Quad BeadChip (Illumina; n = 582,892 SNPs) at the Centre National de Génotypage (Paris, France). Second, the remaining 427 adolescents and all parents were genotyped with the HumanOmniExpress BeadChip (Illumina; n = 729,295 SNPs) at the Genome Analysis Centre of Helmholtz Zentrum München (Munich, Germany). In both genotyping waves, SNPs with call rate <95% and minor allele frequency <0.01 and SNPs that were not in Hardy–Weinberg equilibrium ($P < 1 \times 10^{-6}$) were excluded. After this quality control, 542 345 SNPs on the first chip and 644 283 SNPs on the second chip were available for analysis. Genotype imputation was used to equate the set of SNPs genotyped on each platform and to increase the SNP density. Haplotype phasing was performed with SHAPEIT using an overlapping subset of 313,653 post-quality-control SNPs that were present on both genotyping platforms and the 1000 Genomes SNPs in European reference panel (Phase 1, Release 3). Imputation was conducted on the phased data with IMPUTE2. Markers with low imputation quality (information score <0.5) or low minor allele frequency (<0.01) were removed. After this quality control of imputation, a total of 7,746,837 typed and imputed SNPs were analyzed.

Ethics Statement

Written consents of adults and assent of adolescents (and consent of their parents) were obtained. The regional research ethics committees approved the study protocols.

Funding
The Saguenay Youth Study has been funded by the Canadian Institutes of Health Research (T.P., Z.P.), Heart and Stroke Foundation of Canada (Z.P.) and the Canadian Foundation for Innovation (Z.P.).

Acknowledgements
We thank all families who took part in the Saguenay Youth Study.

### **Tracking Adolescents' Individual Lives Survey (TRAILS)** | Oldehinkel AJ; Hartman CA | PMID 25431468 18263649

<u>Cohort Description</u>

TRAILS (TRacking Adolescents' Individual Lives Survey) is a prospective cohort study of Dutch adolescents with approximately triennial measurements from age 11 to up until adulthood, which consists of a general population and a clinical cohort. Data collection started in 2001, when 2230 (pre)adolescents were enrolled in the study (response rate 76%, mean age 11.1, SD 0.6, 51% girl). In the present study, data were used from the fourth (retention rate 84%, mean age 19.1, SD 0.6 , 52% females), and fifth (mean age 22.3, SD 0.6,53% female) assessment wave of the population cohort, which ran from October 2008 to September 2010 and January 2012 to December 2013 (T5), respectively. For detailed information on the cohort participants and data collected please see PMID 25431468 and 18263649.

<u>Phenotype Measure</u>

Lifetime and last year cannabis use was assessed at the fifth assessment wave (mean age 22 years), by means of a self-report questionnaire. When participants did not have valid data on the fifth wave, we used responses collected during the fourth wave (mean age 19 years). Lifetime use was assessed by the question: "How often did you use weed (marijuana) or hash during your entire life?", and past-year use by the question "How often did you use weed (marijuana) or hash during you're the past 12 months?". Note that this was a multiple choice question: there were single options for 0-10, but higher values were grouped: 11-19, 20-39, 40+.

<u>Genotyping, QC, Imputation</u>

Genome-wide genotyping was performed by the Illumina Cyto SNP12 v2 array. This data was imputed using IMPUTE2 (HRC pannel) and association analysis was performed with SNPTEST v2.4.1.

<u>Ethics Statement</u>

The TRAILS study was approved by the Dutch Central Committee on Research Involving Human Subjects

<u>Funding</u>

TRAILS has been financially supported by various grants from the Netherlands Organization for Scientific Research NWO (Medical Research Council program grant GB-MW 940-38-011; ZonMW Brainpower grant 100-001-004; ZonMw Risk Behavior and Dependence grants 60-60600-97-118; ZonMw Culture and Health grant 261-98-710; Social Sciences Council medium-sized investment grants GB-MaGW 480-01-006 and GB-MaGW 480-07-001; Social Sciences Council project grants GB-MaGW 452-04-314 and GB-MaGW 452-06-004; NWO large-sized investment grant 175.010.2003.005; NWO Longitudinal Survey and Panel Funding 481-08-013 and 481-11-001; NWO Vici 016.130.002 and 453-16-007/2735; NWO Gravitation 024.001.003), the Dutch Ministry of Justice (WODC), the European Science Foundation (EuroSTRESS project FP-006), the European Research Council (ERC-2017-STG-757364 en ERC-CoG-2015-681466), Biobanking and Biomolecular Resources Research Infrastructure BBMRI-NL (CP 32), the Gratama foundation, the Jan Dekker foundation, the participating universities, and Accare Centre for Child and Adolescent Psychiatry. Statistical analyses were carried out on the Genetic Cluster Computer (http://www.geneticcluster.org), which is financially supported by the Netherlands Scientific Organization (NWO 480-05-003) along with a supplement from the Dutch Brain Foundation.

### UK Biobank (UKB)| PMID: 30305743

Cohort Description

The UK Biobank is a large-scale, long-term health study of about 500,000 UK participants aged 40–69 at recruitment [https://pubmed.ncbi.nlm.nih.gov/30305743/]. It offers extensive genetic and health data, including biological measures, mental health questionnaires and genome-wide genotyping.

Phenotype Measure

As part of online mental health and wellbeing questionnaires, each phenotype was defined based on the criteria below:

- Cannabis ever:

Cannabis ever use was assessed by asking: "Have you taken/used cannabis (marijuana, grass, hash, ganja, blow, draw, skunk, weed, spliff, dope), even if it was a long time ago?" With the following response options: "No (0), Yes, 1-2 times (1), Yes, 3-10 times (1), Yes, 11-100 times (1), Yes, more than 100 times (1)". The average age at interview is 64.1 years (SD=7.9, range=40-82 years).

- Cannabis maximum frequency:

Cannabis maximum frequency was assessed in subjects who have used cannabis at least once during their lifetime by asking: "Considering when you were using/taking cannabis most regularly, how often did you take it?" With the following response options: "Less than once a month (0), once a month or more, but not every week (1), once a week or more, but not every day (2), every day (3)". The average age at interview is 59.7 years (SD=8.3, range=40-81 years).

- Cannabis lifetime frequency:

Cannabis lifetime frequency was assessed in subjects who have used cannabis at least once during their lifetime by asking: "Have you used/taken cannabis (marijuana, grass, hash, ganja, blow, draw, skunk, weed, spliff, dope), even if it was a long time ago?" With the following response options: "Yes, 1-2 times (1), Yes, 3-10 times (2), Yes, 11-100 times (3), Yes, more than 100 times (4)". The average age at interview is 61.2 years (SD=8.2, range=40-82 years).

Genotyping, QC, Imputation

We used genotype data from the UK Biobank that had undergone quality control (QC) and imputation [https://pubmed.ncbi.nlm.nih.gov/30305743/]. Individuals who had withdrawn consent or were not classified as having white British genetic ancestry by the UK Biobank were excluded from the analysis. Further QC was performed to remove participants with:

1. Potential sex chromosome aneuploidy.
2. Missing genotype call rate > 5%.
3. Heterozygosity rate (for autosomal markers) ± 3 SD from the mean heterozygosity rate across all individuals.

Moreover, further marker QC was performed to exclude markers with:

1. Imputation info score < 0.4.
2. Minor allele frequency (MAF) < 1%.
3. Call rate < 95%.
4. Hardy–Weinberg equilibrium (HWE) p-value < $10^{-6}$ for cannabis frequency and HWE p-value < $10^{-10}$ for cannabis ever phenotype.

Ethics Statement

Ethical approval for UK-Biobank as a research tissue bank has been provided by the North West Multi-centre Research Ethics Committee (MREC).

Funding

UK Biobank was established by the Wellcome Trust medical charity, Medical Research Council, Department of Health, Scottish Government and the Northwest Regional Development Agency. It is a non-profit charity which has been awarded core funding of around £180 Million. It has also had funding from the Welsh Government, British Heart Foundation, Cancer Research UK and Diabetes UK. Core funding continues to be received from the Wellcome Trust, the MRC, and more recently, from Cancer Research UK and NIHR.

### University of California at San Francisco Family Study (UCSF Family Study) | Gizer IR; Wilhelmsen KC | PMID: 15597083

Cohort Description

The University of California San Francisco Family Alcoholism Study is a nationwide study on the genetics of alcoholism and other substance dependence. Probands were sampled from the community through semi-targeted direct mail, a web site, press releases, advertisements and from alumni of treatment centers across the nation. Probands were invited to participate if they met screening criteria for alcohol dependence at some point in their lifetime and had at least one sibling or both parents available to participate in the study. With the permission of the proband, relatives were invited by mail to participate. Probands with stimulant, cocaine, or opioid dependence and those who reported any history of intravenous substance use were excluded. Probands were also excluded if, upon screening, they reported a current or past diagnosis of schizophrenia, bipolar disorder, or other psychiatric illness involving psychotic symptoms, a life-threatening illness, or an inability to speak and read English. The UCSF cohort consists of approximately 2000 participants aged 18–80 years. Detailed information on study participants and gathered data has previously been published (PMID 15597083).

Phenotype Measure

A modified version of the Semi-Structured Assessment for the Genetics of Alcoholism was used to assess alcohol and other substance use phenotypes, including measures of cannabis use. Average participant age at interview is 45.7 years (SD=10.8, range=17-81 years). Two measures assessing frequency of cannabis use were included, “Did you ever use marijuana at least 21 times in a year?” and “Did you ever use marijuana at least once a week for a month or more?”

Genotyping, QC, Imputation

Blood derived deoxyribonucleic acid was sequenced using a low-coverage whole genome sequencing approach and genotyped using an Affymetrix Exome1A chip to assess accuracy of genotype calls from sequence data. Pair-end sequencing was performed on HiSeq2000 sequencers (Illumina, San Diego, CA). Approximately 86% of samples were sequenced at a coverage depth between 2× and 6× (range: 1×–18×) with depth of coverage evenly distributed across the genome. Sequence reads were aligned using blocked multiple-sequence alignment (BMA), and realigned near indels with the Genome Analysis Toolkit (GATK). Variants were called using the LD-aware variant caller Thunder (Li 2011).

Ethics Statement

Institutional review board committees approved all data collection, and participants provided informed consent prior to participation.

Funding

This work was supported by grants from the National Institute of Alcohol Abuse and Alcoholism: F31AA025269 (principal investigator: Jacqueline M. Otto) and the National Institute on Drug Abuse: R01DA030976 (principal investigators: Kirk C. Wilhelmsen, Ian R. Gizer), and the State of California for medical research on alcohol and substance abuse through the University of California at San Francisco and Ernest Gallo Clinic and Research Center to Kirk C. Wilhelmsen.

### **Utrecht Cannabis Cohort** | Boks, P.M. | PMID: 21156050, 32330591

Cohort Description

The Utrecht Cannabis Cohort (CannabisQuest) consists of 1,259 Dutch young adults aged 18–25 who participated in a genetic sub-study between 2006 and 2009. Participants were drawn from a larger online cohort (N = 17,698) recruited via www.cannabisquest.nl, but only those who were assessed at the university hospital provided DNA samples through venipuncture and were included.
Participants were selected in two waves: Wave 1: 719 individuals selected using an extreme sampling strategy based on cannabis use (lifetime use <2 or >10 times) and psychotic-like experiences (top or bottom CAPE quintiles). Wave 2: 540 individuals from an unselected subsample.
Inclusion was limited to individuals with four grandparents born in the Netherlands to reduce genetic heterogeneity. Participants completed online or local assessments. A subset received structured psychiatric interviews (SCID or MINI). Recent cannabis use was validated via urine drug screening.

Phenotype Measure

Lifetime cannabis use (never, 1, 2, 5–9, >10 times)
Weekly cannabis expenditure (THC proxy: <€3, €3–10, €10–25, >€25)

Genotyping, QC, Imputation

- DNA collected from two 10 ml EDTA tubes via venipuncture
- Genotyping platforms:
    - Illumina HumanOmniExpress (n = 576; 733,202 SNPs)
    - Illumina Human610-Quad Beadchip (n = 768; 620,901 SNPs)
- QC procedures (PLINK):
    - Exclusion of participants with sex mismatches or >5% missing genotypes
    - SNP-level filters: MAF >5%, HWE p >1e-6, SNP missingness <2%
    - LD pruning ($R^2 < 0.2$) and merging with HapMap Phase 3 for ethnicity check
- Imputation: HapMap 3 Release 24 using Beagle

Ethics Statement

Approved by the Medical Ethics Committee of the University Medical Center Utrecht. All participants provided written informed consent.

Funding

The Utrecht Cannabis Cohort was funded by NWO, the Dutch council for scientific research (ZonMW TOP grant no. 91207039).

### Virginia Twin Study of Adolescent Behavioral Development (VTSABD) |

PIs: Lindon Eaves, Judy Silberg, Hermine Maes | PMID: 9294370

Cohort Description

The Virginia Twin Study of Adolescent Behavioral Development (VTSABD) is a population-based multi-wave cohort-sequential study of twins born between 1974 and 1983, ascertained primarily through the Virginia state school system and participating private schools. It serves as one of the four sites in the Gene-Environment-Development Initiative (GEDI). Of 1894 putative twin pairs, 1412 families (75%, 2775 children) participated and were included in the first wave of data collection. Three subsequent waves of data collection occurred at approximately 1½-year intervals, and a fifth wave when participants were in their mid-20s, and a sixth wave when participants were in their late-20s. The study was limited to subjects of European ancestry as insufficient numbers from other ancestry groups were ascertainable. Parents completed similar assessments for both twins. After age 18, the twins alone were interviewed individually by telephone or questionnaire. Over 10,000 individual assessments have been completed, with 2289 (82%) of the Wave 1 sample completing the fifth wave.

Phenotype Measure

As part of a comprehensive psychiatric assessment using the Child and Adolescent Psychiatric Assessment (CAPA) and Life Experiences Interview (LEI), the VTSABD collected detailed information on marijuana use patterns. For participants who reported ever using marijuana or hashish, the study assessed numerous aspects including: age of onset of first use, whether use occurred again soon after first use, regularity of use, current use status, problems associated with use, frequency of use per month, and whether use exceeded 10 times per month. Additionally, the study assessed multiple diagnostic criteria for marijuana abuse and dependence based on DSM-IV, including failure to fulfill obligations, hazardous use, interpersonal problems, legal problems, tolerance, withdrawal symptoms, using more than planned, quit attempts, giving up activities, time spent using, and psychological/ physical problems resulting from use.

Genotyping, QC, Imputation

Nine ml. of blood were collected from VTSABD participants in the first year of the GEDI study, when subjects were aged 25 to 34. Blood and informed consent for genotyping and storage in dbGaP were obtained from 913 participants, of whom 281 were co-twins. The blood samples were sent to the Rutgers University Cell and DNA Repository for DNA extraction, and to the Genotyping Shared Resource at the Mayo Clinic Cancer Center for genotyping. All samples were genotyped using Illumina Human660W-Quad v1 DNA Analysis BeadChips. Quality control was carried out in the Department of Genetics at the University of North Carolina, Chapel Hill.

Ethics Statement

The Institutional Review Boards at all data gathering sites approved this study, and written consent was obtained from all participants.

Funding

The VTSABD/GEDI was supported by the National Institute on Drug Abuse (NIDA) as part of the Gene-Environment-Development Initiative (GEDI) under grant R01DA024413 and by the National Institute of Mental Health under grant R01MH45268.
